# Bead-based approaches to CRISPR diagnostics

**DOI:** 10.1101/2023.09.03.23294926

**Authors:** Sameed M. Siddiqui, Nicole L. Welch, Tien G. Nguyen, Amaya Razmi, Tianyi Chang, Rebecca Senft, Jon Arizti-Sanz, Marzieh E. Mirhashemi, David R. Stirling, Cheri M. Ackerman, Beth A. Cimini, Paul C. Blainey, Pardis C. Sabeti, Cameron Myhrvold

## Abstract

CRISPR-based diagnostics have emerged as a promising tool for fast, accurate, and portable pathogen detection. There has been rapid progress in areas such as pre-amplification processes and CRISPR-related enzymes, but the development of reporter systems and reaction platforms has lagged behind. In this paper, we develop new bead-based techniques that can help fill both gaps. First, we develop a novel bead-based split-luciferase reporter system with improved sensitivity compared to standard fluorescence-based reporter design in CRISPR diagnostics. Second, we develop a highly deployable, bead-based platform capable of detecting nine distinct viral targets in parallelized, droplet-based reactions. We demonstrate the enhanced performance of both approaches on synthetic and clinical samples. Together, these systems represent new modalities in CRISPR diagnostics with increased sensitivity, speed, multiplexing, and deployability.

## Introduction

The COVID-19 pandemic has highlighted the need for rapidly deployable diagnostic technologies able to respond to new pathogens and emergent variants anywhere in the world (Liu et al. 2021). However, no current technology uniquely meets the sensitivity, specificity, deployability, speed, and multiplexing needs required for a broad and robust response to infectious disease outbreaks. Quantitative polymerase chain replication (qPCR), widely considered a gold standard due to its sensitivity and specificity, cannot be deployed readily, and remains limited in multiplexing ability (Eckbo et al. 2021; Center for Devices and Radiological Health n.d.; Knox and Beddoe 2021). Next generation sequencing (NGS) is similarly sensitive, specific, and able to detect many pathogens and variants; however, it is expensive, has a long turnaround time, and requires significant technical expertise to deploy for viral surveillance (Goodwin, McPherson, and McCombie 2016). On the other hand, antigen capture tests are readily deployable and affordable, but are less sensitive and specific than nucleic acid tests and are not rapidly adaptable for new pathogens (Arizti-Sanz et al. 2020; Chu et al. 2022).

CRISPR-based systems offer an alternative approach that is well-poised to address pathogen diagnostic needs. Specifically, CRISPR effectors Cas12 and Cas13 exhibit collateral cleavage activity upon recognition of their target DNA or RNA, respectively, enabling these enzymes to act as target-specific sensors (Chen et al. 2018; Liu et al. 2021; Gootenberg et al. 2017; East-Seletsky et al. 2016). Since their introduction, there has been substantial work in developing CRISPR-based diagnostic systems, with assays for diverse pathogens such as influenza, Zika virus, and SARS-CoV-2 developed across different combinations of CRISPR effectors, imaging tools, and reaction platforms (Pardee et al. 2016; Liu et al. 2021; Park et al. 2021; Fozouni et al. 2021). Much of this effort has been limited to fluorescence and lateral flow strip readouts, leaving room for orthogonal advances in reporter design and reaction barcoding to improve deployability, sensitivity, and multiplexing capability (Myhrvold et al. 2018; Barnes et al. 2020; Welch et al. 2022; Arizti-Sanz et al. 2020; Ackerman et al. 2020; Arizti-Sanz et al. 2022; Liu et al. 2021).

New bead-based systems have led to recent advances in protein detection due to their ability to compartmentalize reaction components and may serve as a basis to advance CRISPR-based nucleic acid detection. For example, AlphaLISA implements a two-bead chemiluminescent reporter system that enables highly-sensitive wash-free antigen detection (Ullman et al. 1994; Bielefeld-Sevigny 2009). The two beads in the system have different surface chemistries and are coupled with complementary antibodies of a sandwich ELISA; when an antigen is bound, the complementary beads come together and emit light as a reporter for antigen detection. This suggests the possibility of a highly sensitive split luciferase reporter for Cas13 diagnostics which separates split luciferase components onto different beads prior to Cas13 target detection. Separately for multiplex detection, Luminex consists of fluorescently color-coded beads that are coupled to different antibodies, enabling pooled identification of separate targets in a single reaction (Fulton et al. 1997; Anderson et al. 2011). This technology suggests that color-coded beads could lend themselves well to having different targets for Cas13 detection on each bead. We explored both technological approaches to expand the breadth of CRISPR-based diagnostic platforms.

We first considered how bead-based readouts could improve sensitivity in point-of-need CRISPR-based diagnostic assays, such as in Streamlined Highlighting of Infections to Navigate Epidemics (SHINE), a point-of-need CRISPR diagnostic platform which increases sensitivity by coupling isothermal amplification with Cas13 detection (Arizti-Sanz et al. 2022, 2020). These assays have traditionally used fluorescence-based reporters, primarily consisting of a fluorescein (FAM) dye linked by a short oligonucleotide sequence to a quencher (Arizti-Sanz et al. 2020; Chen et al. 2018; Liu et al. 2021; Myhrvold et al. 2018). While these assays have performed well, fluorescence-based technologies are known to have high background signal and low sensitivity compared to bioluminescence technologies (Arizti-Sanz et al. 2020; Tung et al. 2016).

A bead-based luminescent split reporter system which links nucleic acid detection with NanoLuciferase (NanoLuc) complementation could provide an attractive alternative to fluorescent reporters, enabling rapid attomolar detection with a high dynamic range (Dixon et al. 2016; Schwinn et al. 2018; Fan and Wood 2007). A two-bead system with a large protein subunit (LgBiT) and a smaller peptide subunit (HiBiT) each coupled to separate bead type may serve as a basis for a Cas13 cleavage reporter if at least one of the protein subunits is coupled via a Cas13-cleavable RNA linker. By virtue of being coupled to separate beads, LgBiT and HiBiT can be largely separated from each other and kept catalytically inactive. In the presence of the target, Cas13 collateral cleavage of the bead RNA linkers could reverse this separation and allow the formation of complemented NanoLuc. Therefore, for point-of-need use, a new split-luciferase-based reporter system could be well-poised to improve sensitivity in amplification-free conditions while simultaneously removing the requirement of a light source as in fluorescence-based systems.

We next considered how a bead-based system could improve multiplexed diagnostic testing at point of care. We previously developed the CRISPR-based Combinatorial Arrayed Reactions for Multiplexed Evaluation of Nucleic acids (CARMEN) and microfluidic CARMEN (mCARMEN) (Welch et al. 2022; Ackerman et al. 2020), and demonstrated their unprecedented multiplexing and sensitivity across samples and pathogens. These platforms, however, require high technical expertise and costly equipment to achieve sample or patient barcoding, restricting their deployability in resource-limited settings (Ackerman et al. 2020; Welch et al. 2022). This constraint leaves an opportunity to replace barcoding with a less resource-intensive, bead-based approach.

A color-coded bead-based approach that couples beads to distinct crRNAs could be used to create a localized separation of crRNA, enabling an assay with multiplexed nucleic acid targets. Given previous work in microparticle-based dropletization, such color-coded beads could allow equipment-free nanoliter droplet generation (Clark et al. 2023), with each droplet containing Cas13 detection master mix and approximately one color-coded crRNA bead. With crRNA-specific detection reactions occurring in parallel across different droplets, this could in turn enable a highly parallelized reaction which could be imaged in a fluorescent microscope or plate reader to determine bead target-specific detection. Thus, for point-of-care use, a bead-based, low-cost platform capable of parallelized, dropletized detection of multiple targets may enable a sensitive, robust, and highly multiplexed solution for resource-limited settings.

Here we explore the applicability of novel bead-based approaches to increasing sensitivity, multiplexing, and deployability in CRISPR-diagnostics. First, we designed a bead-based split luciferase reporter (bbLuc) and examined this readout modality in an amplification-free reaction and in the SHINE diagnostic platform. Next, we developed and validated a bead-based deployable multiplexed diagnostics platform (bbCARMEN) and further investigated its performance in an implementation of a panel of nine respiratory viruses (Welch et al. 2022).

## Results

### Bead-based strategy to couple Cas-13 activity with a split NanoLuciferase-based readout

We developed a bead-based Luciferase reporter (bbLuc) to couple Cas13 activity with a split Nanoluciferase-based readout (Figure 1A). As a first step, we probed whether it is possible to couple HiBiT and LgBIT to beads using a Cas13-cleavable RNA-based linker. We attached HiBiT and LgBiT to biotinylated oligonucleotides using HaloLigand-HaloTag-based covalent linking, thereby enabling coupling to streptavidin coated beads (Figure 1B) (Los et al. 2008). Notably, this coupling enabled target-mediated Cas13 cleavage of HiBiT-linked nanoparticles (Figure 1C). However, we did not observe significant Cas13 cleavage of LgBiT-linked nanoparticles, most likely due to increased steric hindrance of Cas13 cleavage in proximity with the larger LgBiT enzyme compared to the HiBiT peptide. As such, we focused our subsequent design efforts on cleavable HiBiT-nanoparticles.

**Figure 1:**
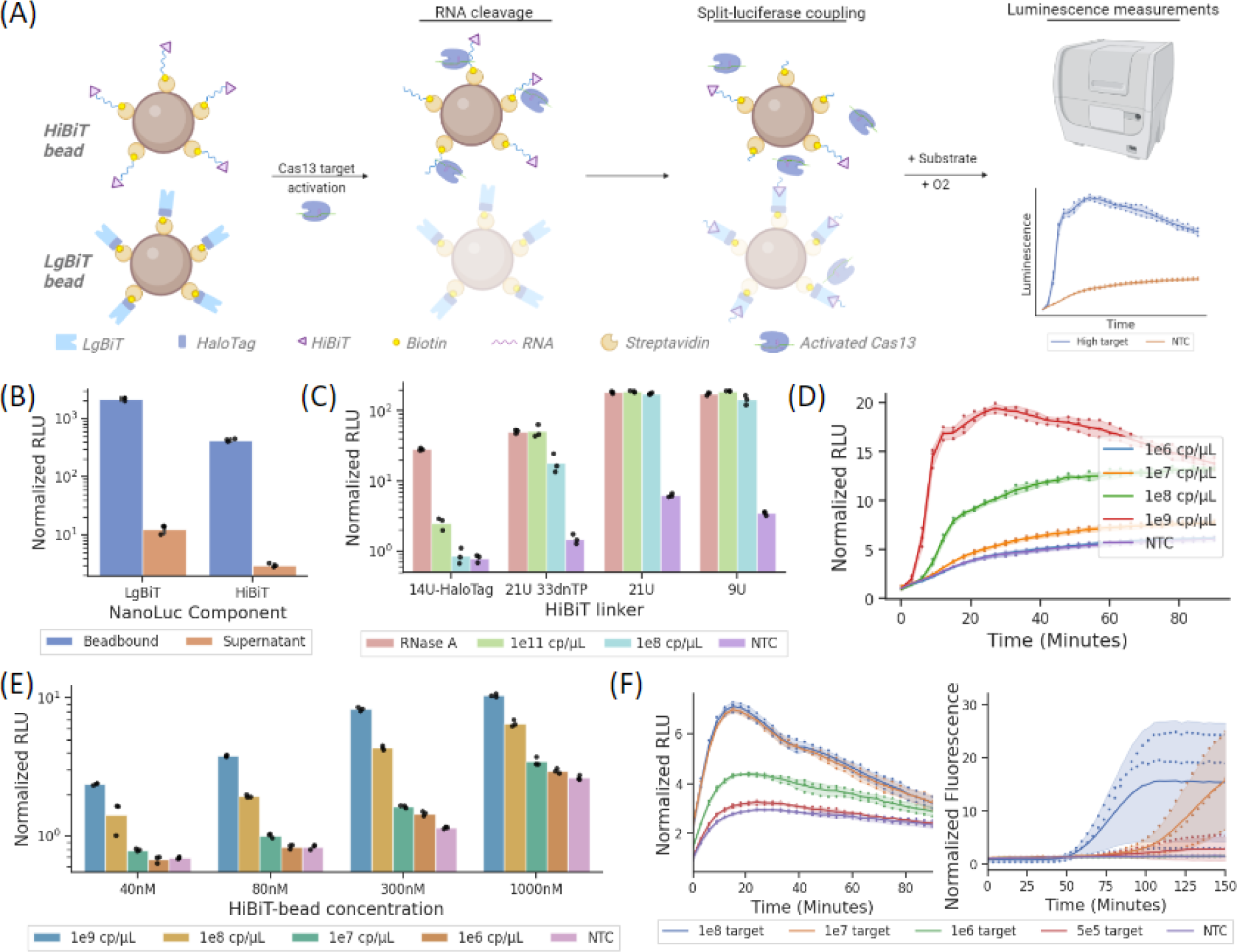
Initial development for bbLuc. (A) Schematic of split-luciferase reporter system. HiBiT and LgBiT are separated before Cas13 conjugation via their coupling to different beads to enable localized separation. (B) Beads bound with 80 nM HiBiT or 80 nM LgBiT show durable coupling through storage and wash cycles. Luminescence values normalized to the average NTC signal at the first collected timepoint. (C) One-bead luminescent assay to compare different RNA linkers used to couple HiBiT to beads (21U 33dnTP HaloLigand-HaloTag based, 21U SPAAC-based, 9U SPAAC-based linkers). RNase A or Cas13/target mixes were added to coupled beads to enable cleavage, with the resulting cleaved products separated from the beads magnetically and added to LgBiT in solution. (D) Luminescent kinetics of full assay, prior to bead optimization (80 nM HiBiT, 80 nM LgBiT). The normalized signal is lower than in previous reactions due to differences in on-bead versus in-solution reaction kinetics. (E) Optimization of HiBiT bead concentrations in amplification-free luminescent assay. LgBiT concentration maintained at 80 nM on varied synthetic RNA target; NTC, no target control. (F) Optimized full luminescent detection assay kinetics compared to fluorescent assay on varied synthetic RNA target.

Having demonstrated the ability to couple HiBiT to beads using a Cas13-cleavable RNA-based linker, we next designed improved linkers to optimize and enable more efficient cleavage. We found through a side-by-side comparison of Cas13-based and RNase A-based cleavage that the initial Cas13 cleavage of HaloTag-based linking was inefficient (Figure 1C). We hypothesized that this inefficiency may have been caused again by steric hindrance between the beads and HaloTag-HiBiT complex, reducing accessibility of Cas13 to cleavage sites. We therefore increased linker length (Supplemental Table 1) and changed linkage chemistry by using Strain-promoted Azide - Alkyne Click Chemistry reaction (SPAAC) to connect the HiBiT peptide to the oligonucleotide linker. This linkage enabled significantly more efficient Cas13 cleavage compared to the HaloTag-HiBiT, showing equivalent cleavage of the HiBiT linker from Cas13 and RNase A (Figure 1C).

Next, we characterized and further optimized the performance of an amplification-free assay with both HiBiT and LgBiT beads in solution. Using 80nM HiBiT peptide coupled to nanoparticles in solution, we were able to achieve detection down to 10^7^ copies/uL of input RNA (Figure 1D). While this compared favorably to a fluorescence-based amplification-free assay, we observed further improvements in sensitivity when the surface density of HiBiT peptides on nanoparticles was increased to a concentration of 300nM (Figure 1E). HiBiT concentrations above 300nM and LgBiT concentrations above 80nM resulted in more inconsistency and lower sensitivity (Supplementary Figure 2), possibly due to an increased viscosity of the solution caused by a higher peptidic charge on the beads.

We compared the performance of bbLuc to a conventional fluorescent reporter in the amplification-free assay. Our luminescent reporter detected down to 5*10^5^ copies/µL of input target compared to 1*10^7^ copies/µL for the fluorescent reporter. We were thus able to achieve a 20x increase in sensitivity using the luminescent reporter (Figure 1F, 2A).

### Integration of luminescent reporter into SHINE

Having optimized bbLuc in an amplification-free setting, we then assessed its performance in the diagnostic platform of SHINE. We found that the luminescent reporter performed 5x better than the conventional fluorescent reporter in the SHINE setting, compared to the 20x enhancement we achieved in the amplification-free setting (Figure 2A). We hypothesized that the greater enzymatic and buffer complexity of SHINE compared to amplification-free assays may have allowed for deleterious interactions between the bead complexes and reaction mixture, causing the observed 4x decrease in relative sensitivity.

**Figure 2:**
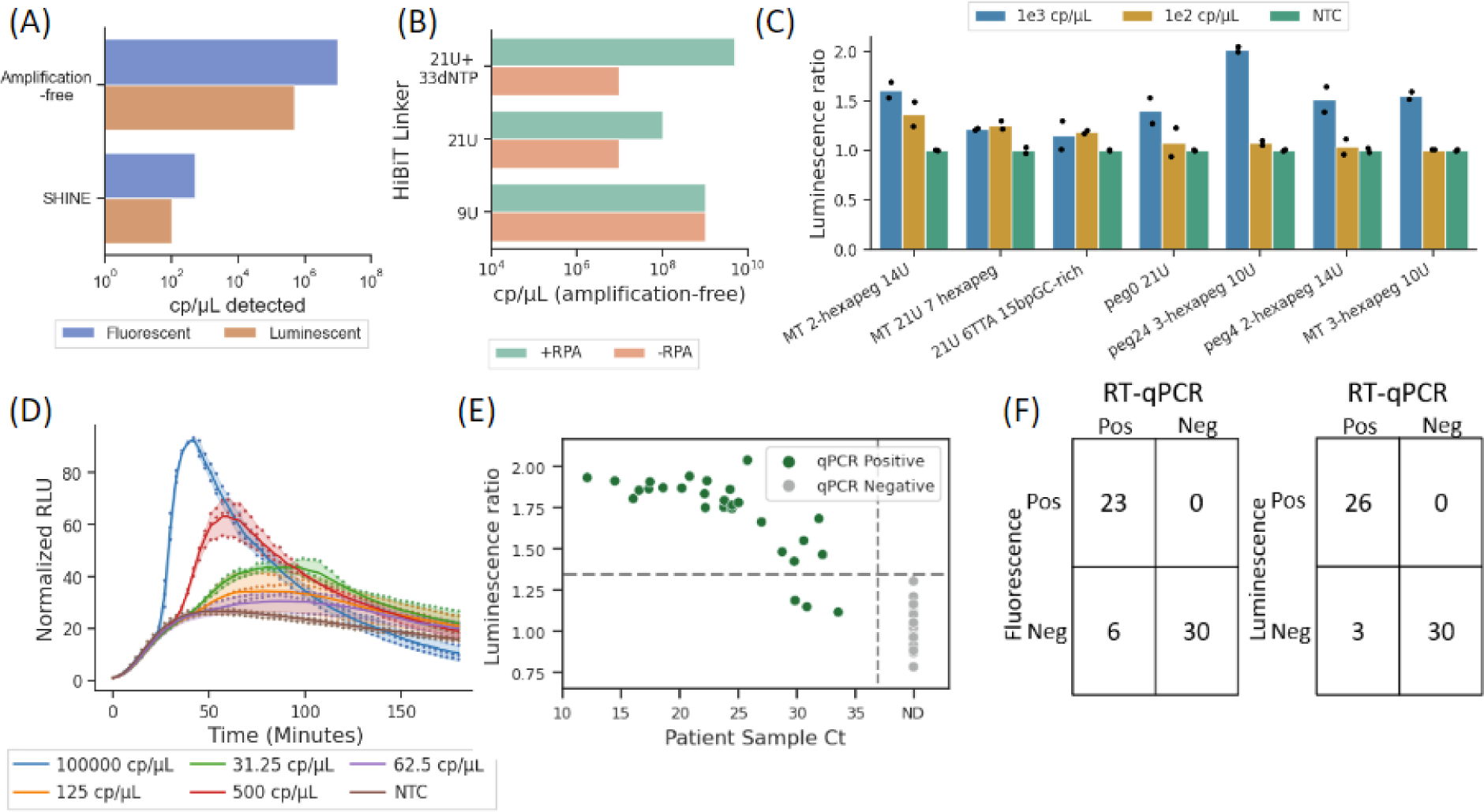
bbLuc integration with SHINE. (A) Luminescence comparison of SHINE and amplification-free systems prior to optimization. Luciferase amplification-free and SHINE data collected after 30 minutes and 90 minutes, respectively; fluorescence amplification-free and SHINE data collected after 3 hours. (B) Luminescent SHINE results of beads with different RNA linkers shows differential effects after 3h with and without recombinase polymerase (RPA) (80 nM LgBiT, 300 nM HiBiT) (C) Luminescent SHINE performance of 22 tested RNA linkers after 3h (80 nM LgBiT, 300 nM HiBiT) (D) Final, optimized reaction kinetics with luminescent SHINE (80 nM LgBiT, 300 nM HiBiT, 2-hexapeg 14U linker, 80nM furimazine, 140nM RPA primers) (E) SARS CoV-2 detection using RNA from patient samples (80 nM LgBiT, 300 nM HiBiT). Threshold line plotted as 1.35, determined empirically. (F) SARS CoV-2 detection of detected patient samples using luminescent and fluorescent SHINE compared to RT-qPCR. A-D on varied synthetic RNA target

We explored the potential causes of reduced sensitivity of our luminescent reporter in the SHINE context by testing different concentrations of a molecular crowding reagent and assessing if individually spiking in SHINE components to amplification-free reactions interfered with detection. We first examined possible molecular crowding effects caused by bead addition to the assay by modifying polyethylene glycol (PEG) concentration and molecular weight in the SHINE reaction buffer, but found the PEG parameters in fluorescent SHINE were optimal with the luminescent system (Supplementary Figure 3). We then probed possible inhibitory effects on the beads from the constituent SHINE proteins by individually spiking reaction components into separate amplification-free reactions, finding a 1000-fold reduction in performance upon addition of recombinase polymerase amplification (RPA) pellets (Supplementary Figure 4). We found that this reduction in sensitivity was largely caused by single-stranded binding (SSB) protein reducing Cas13 cleavage of the beads (Supplementary Figure 5).

Based on the observation that the SSB used in RPA was inhibiting our bead linkers, we tested a series of new linkers to find designs that improved detection in SHINE. We hypothesized that our original HiBiT-bead linker design, consisting of 54 nucleotides (21 uracils and 33 dNTPs) may have served as a binding site for SSB, thereby interfering with Cas13 cleavage of the linker. We first tested shorter linker designs without any dNTPs, finding reduced inhibition of detection by 21U and 9U linkers in the presence of RPA components compared to our original design (Figure 2B). We went on to test 27 different linker designs (Figure 2C, Supplementary Figures 6-7, Supplemental Table 1), finding that a 2-hexapeg 14U linker provided the highest sensitivity amongst our reporters.

We made a few final optimizations and measured the limit of detection (LOD) of our assay. We varied the concentrations of furimazine, RPA primers, and magnesium acetate, determining optimal concentrations of 80nM, 140nM, and 14nM, respectively (Supplementary Figures 8-10). We found our optimized assay had an LOD of 32 copies/uL of input RNA in 75 minutes (Figure 2D), representing a 3x improvement of sensitivity following optimization.

### SARS-CoV-2 clinical sample validation of luminescent reporters

We compared the performance of bbLuc and fluorescent SHINE to a gold-standard RT-qPCR on RNA extracted from 63 clinical swabs from suspected COVID-19 patients. Four out of 63 samples were negative for RNase P, a control gene that confirms adequate sample collection. Among the remaining 59 samples, 29 were RT-qPCR confirmed COVID-19 positive and 30 were confirmed COVID-19 negative. The remaining four were ruled inconclusive as they were negative for the Rnase P control test. bbLuc SHINE detected SARS-CoV-2 in 26 of the 29 positive samples (89.6% concurrence) compared to 23 of the 29 positive samples (79.3%) in fluorescent SHINE. Every positive sample detected by fluorescent SHINE was also detected as positive by our luminescent system, which additionally detected 3 high Ct (Ct > 28.5) samples that the fluorescent SHINE did not. This demonstrates an increased sensitivity with luminescent reporters. Both fluorescent and luminescent SHINE correctly identified all 30 RT-qPCR negative samples as negative.

### Equipment-free bead-based droplet generation for multiplexed fluorescent Cas13 detection

Next, we developed a bead-based approach for localizing separation of reaction components to conduct discrete, multiplexed testing in the same overall solutions for point-of-care settings. To enable bead-based multiplexing of target detection, we first attached target-specific biotinylated crRNAs to color-coded, streptavidin-coated beads, examining results when cRNA was attached to beads via either 3’ end biotinylation or via 5’ end biotinylation (Figure 3A, Supplementary Figure 11). With 3’ end biotinylation, we found significant Cas13 cleavage in target compared to no target control (NTC) down to 1 copy/uL of input. In comparison, with 5’ end biotinylation, Cas13 cleavage was not able to distinguish an input of 10^4^ copies per microliter from NTC (Supplementary Figure 12). As such, we used 3’ biotinylated crRNA beads moving forward.

**Figure 3.**
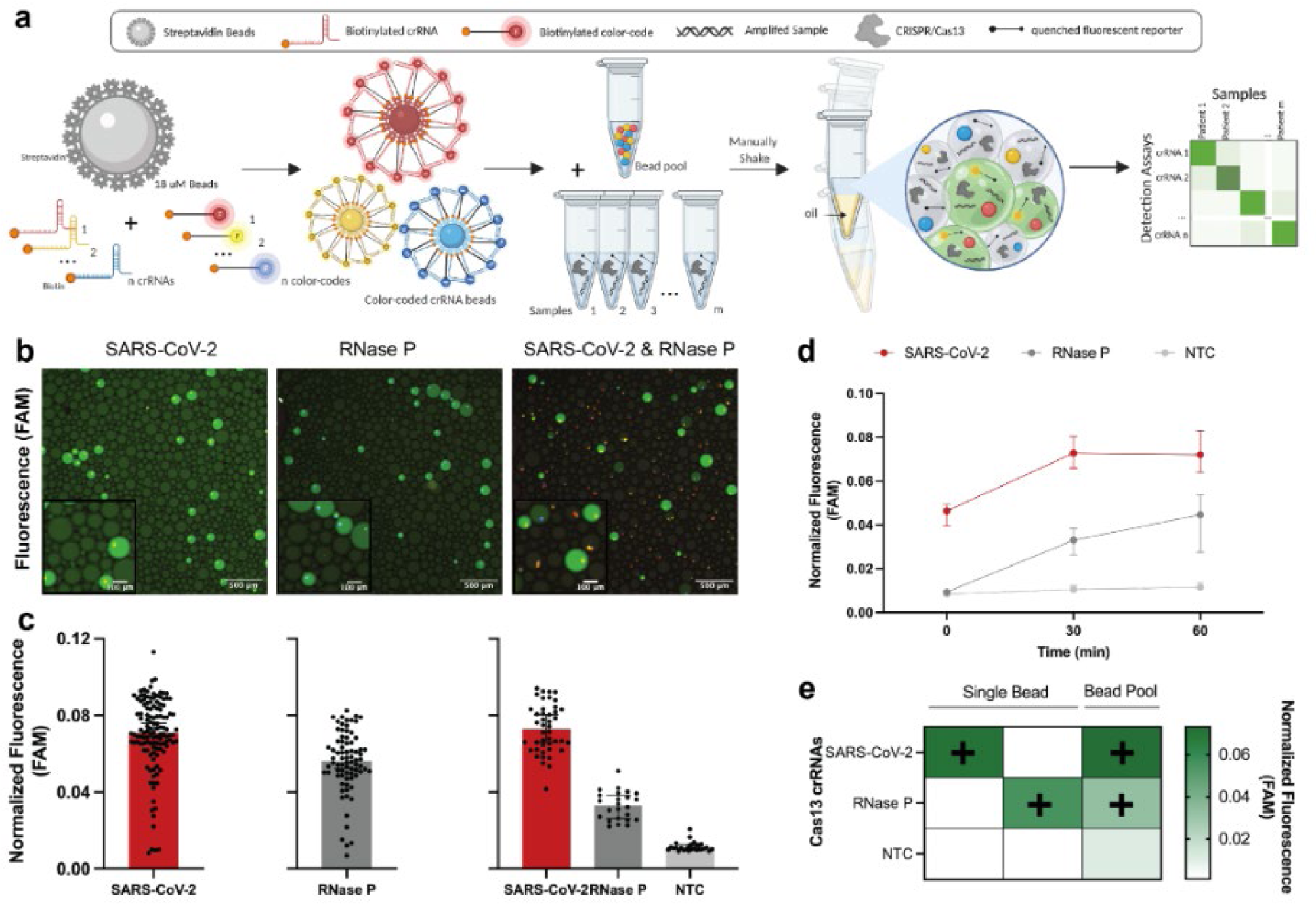
Development of bbCARMEN with biotinylated crRNA-beads in equipment-free generated droplets. **a**, Schematic of bbCARMEN workflow. **b,** Merged fluorescent images of color-coded crRNA beads in droplets at 500 um. SARS-CoV-2: AF546 shown as yellow; RNase P: AF647 shown as blue; FAM reporter: shown as green. **c,** Fluorescence from droplets from individual crRNA bead solutions and pooled bead solutions at 30 min post-reaction initiation. Individual points represent fluorescence from individual droplets. Median fluorescence with 95% confidence intervals (CIs). **d,** Fluorescence kinetics of amplified SARS-CoV-2 synthetic gene fragment at 106 copies/uL and RNase P in pooled crRNA bead solution from b & c. Red: SARS-CoV-2, Dark Gray: RNase P; Light Gray: NTC. Median fluorescence with 95% confidence intervals (CIs). **e**, Heatmap showing SARS-CoV-2, RNase P, and NTC median fluorescence values at 30 min post-reaction initiation in single and pooled crRNA bead solutions from b & c. NTC: no target control.

To show that our bead-based CARMEN (bbCARMEN) approach would work in the context of multiple targets, we developed assays for SARS-CoV-2 and a human internal control RNAse P both individually and in combination (Figures 3B-E, Supplementary Figure 13). We added color-coded beads to a solution containing Cas13 and additional detection components with amplified patient samples. For each reaction, we combined the detection master mix with oil, shaking to form miniature droplets containing the master mix and one color-coded bead (Figure 3B, Supplementary Figure 12). We then loaded samples on custom, prefabricated flow cells for readout using fluorescence microscopy and automated image analysis to track both the color-coded crRNA beads and the signal from Cas13 activity (Figure 3C-D, Figure 4A, Supplementary Figures 12-13). By 30 minutes, we were successfully able to detect fluorescence intensity in droplets above background for all conditions (Figures 3D-E, Supplementary Figure 13).

**Figure 4.**
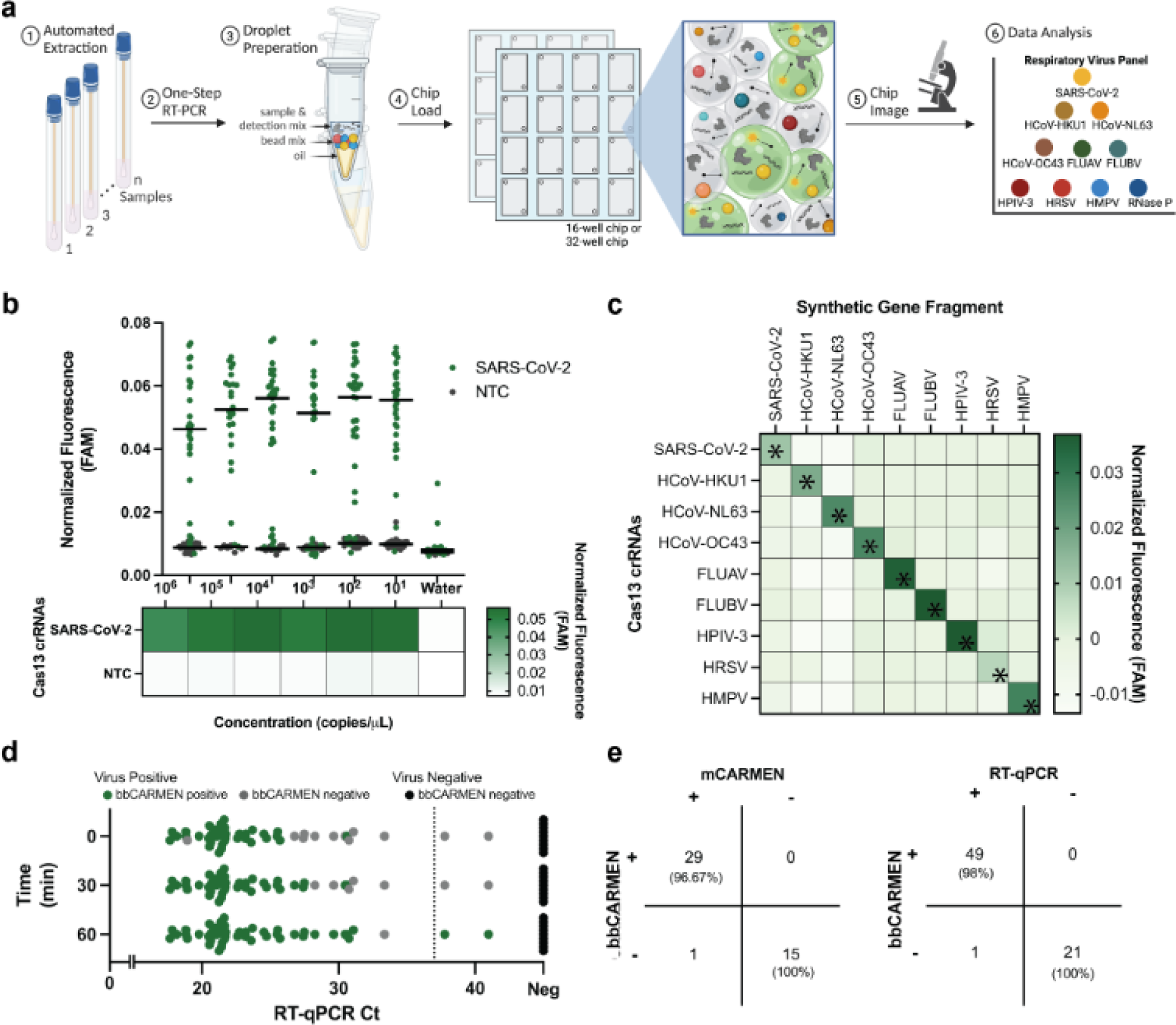
Implementation of Respiratory Virus Panel (RVP) on bbCARMEN. **a**, Schematic of multiplexed RVP assay with each of the 9 viruses on panel differentiated by distinct color-codes. **b,** Fluorescence across SARS-CoV-2 dilution series from 10^6^-10^1^ copies/uL and corresponding NTC fluorescence in a pooled crRNA bead solution. Green: SARS-CoV-2; Gray: NTC. Bar at median fluorescence 30 min post-reaction initiation. **c,** Fluorescence at 10^4^ copies/uL for all 9 viruses on RVP. Fluorescence shown as the median value across 20 replicates at 30 min post-reaction initiation. **d,** Scatter plot comparison of SARS-CoV-detection at 0, 30, and 60 min time points using bbCARMEN and RT-qPCR. **e**, Concordance of RT-qPCR and NGS results compared to CARMEN v2. Right, RT-qPCR results. Left, NGS results.

### Respiratory virus panel implementation on bbCARMEN

We next tested the performance of a larger multiplexed bbCARMEN by implementing a 9-target Respiratory Virus Panel (RVP) previously characterized on the mCARMEN platform (Figure 4A) (Welch et al. 2022). We conjugated human RNase P crRNA (as an internal control) and each crRNA from the RVP to beads with target-specific color-codes and added the beads to the reaction along with patient sample (Supplementary Figure 14). We used fluorescence microscopy to track patient sample signals over time and map crRNA/color-code combinations (Supplementary Figure 14).

With the RVP, bbCARMEN successfully distinguished all bead color-codes from one another across replicates, while simultaneously observing on-target signal with minimal-to-no off-target signal for each panel member (Figures 4B-C, Supplementary Figure 14). We verified the accuracy of these results by conducting LOD studies for all panel members (Figure 4B, Supplementary Figures 15-16). Overall, we found the LOD for each virus (SARS-CoV-2 and HCoV-HKU1: 2,500 copies/mL: FLUBV and HPIV3: 5,000 copies/mL: FLUAV and HRSV: 10,000 copies/mL, HMPV: 20,000 copies/mL;HCoV-NL63 and HCoV-OC43: 40,000 copies/mL) to be in line with or slightly higher than the LOD of the RVP on mCARMEN.

To more rigorously validate sensitivity, we tested 47 freshly collected (Delta and Omicron) SARS-CoV-2 specimens and 9 negatives based on RT-qPCR and NGS results (Figure 4D-E). All but one positive specimen (97.9%) was deemed virus-positive by bbCARMEN within 60 minutes, and all negative samples were correctly deemed negative. To further assess the sensitivity of this assay, we also tested bbCARMEN on two cohorts of virus-positive samples previously characterized with mCARMEN and clinically-validated comparator assays and stored in −80C (Supplementary Figure 17). Of the 60 positive samples tested (30 SARS-CoV-2 positive and 30 HSRV positive), bbCARMEN detected 56 (93.33%) within 60 minutes, exhibiting robust sensitivity even in samples subjected to a freeze-thaw cycle.

### Automated readout of bbCARMEN using commercially available consumables and equipment

To further simplify deployment of this assay in low-resource settings, we reconfigured bead loading and imaging steps to use a standard well plate and a lab plate reader instead of our previous custom flow cells and fluorescent microscope setup (Figure 5A). We found that 96-well plates loaded with a thin layer of droplets contained too many overlapping droplets of each color code for statistical significance in the final diagnostic calls. However, loading onto 48 well plates enabled the droplets to be spread out over a larger area, enabling us to distinguish between different color codes.

**Figure 5.**
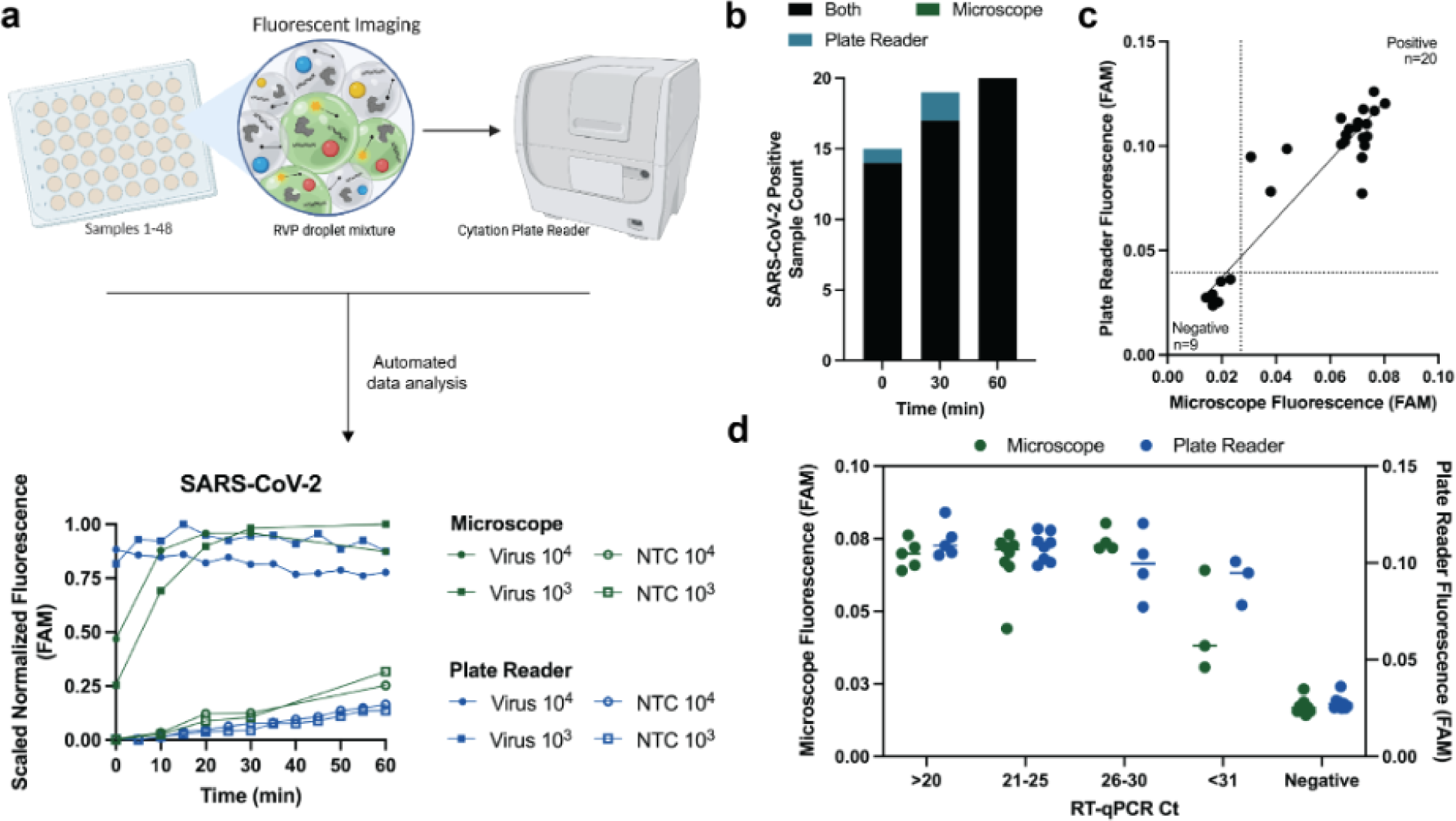
Automated readout of bbCARMEN reduces hands on time and customized equipment requirements. a, Schematic of plate-reader platform, including comparison of bbCARMEN detection kinetics on the two readout platforms. **b,** bbCARMEN SARS-CoV-2 positive hit call at time 0, 30, and 60 mins. Green: positive only by microscope. Blue: positive by plate reader only. Black: positive by both. **c,** Comparison of fluorescent signal using synthetic targets. Thresholds for positivity shown as dashed lines. **d,** SARS-CoV-2 Patient sample detection compared between readout platforms.

We compared concordance of assay results between the microscopy-based and plate reader and found 100% concordance between the two readout platforms across synthetic material and patient specimens (Figures 5B-D, Supplementary Figure 18). Moreover, the plate reader method showed earlier discrimination between target conditions and NTC than our previous microscopy approach, suggesting an improvement in both assay workflow and performance (Supplementary Figure 19). As such, the changes to bead loading and imaging reduce equipment requirements, labor intensity, and data analysis expertise while maintaining sensitivity and sensitivity of bbCARMEN.

## Discussion

In this study, we employ novel bead-based approaches to achieve point-of-need and point-of-care uses in CRISPR diagnostics. These technologies increase sensitivity and deployability in resource-constrained point-of-care settings.

Our luminescent bead-based approach, bbLuc, provides an attractive alternative to traditional fluorescence-based diagnostics, showing increased sensitivity in synthetic and clinical specimens. Critically, enhanced sensitivity also has upstream effects on assay adaptation to new or emerging pathogens by reducing the optimization time required to meet a target LOD. Furthermore, a luminescent assay reduces equipment requirements in conventional assays by removing the need for a light source for fluorescence excitation.

By utilizing a multiplexed bead-based system for point-of-care diagnostics, bbCARMEN addresses the significant equipment and expertise requirements of other multiplexed systems (Welch et al. 2022; Ackerman et al. 2020). bbCARMEN maintains excellent multiplexing ability and sensitivity by using beads as an operationally simple, inexpensive modality to perform multiplexed reactions with high specificity as shown in our clinical sample testing. Implementation of our viral respiratory panel assay further demonstrates the ease of adaptability and its potential to dramatically increase deployability in resource-constrained settings.

There are numerous avenues to enhance both bead-based technologies. For example, in bbLuc, we focused on the cleavage of HiBiT-nanoparticles instead of LgBiT-nanoparticles. Due to manufacturing constraints, we were not able to consistently manufacture an RNA linker long enough for Cas13 cleavage of LgBiT nanoparticles, but future work may incorporate new technologies in RNA synthesis or protein-oligo conjugation. In bbCARMEN, we were able to successfully resolve 9 different crRNA color-codes, giving resolution to discriminate against different viral infections in a point-of-care setting. However, future work can be undertaken to improve the number of simultaneously assayable viruses by iterating on color code technology, either through the use of new fluorescent dyes, a different combinatorial barcoding strategy, or novel multicolor bead approaches.

The fundamental advances of these bead-based platforms may be applied in the future to other CRISPR-based diagnostic platforms. Together, these technologies represent new modalities to increase diagnostic sensitivity and portability, opening avenues for the rapid and sensitive detection of biological molecules.

## Methods

### Clinical samples and ethics statement

The use of excess human specimens, including nasopharyngeal swabs from Boca Biolistics, for use by the Broad Institute were reviewed and approved by the MIT Institutional Review Board under protocol no. 1612793224. Human specimens from patients with SARS-CoV-2, HCoV-HKU1, HCoV-NL63, FLUAV, FLUBV, HRSV and HMPV were obtained under a waiver of consent from the Mass General Brigham IRB protocol no. 2019P003305.

### Sample collection and extraction

Patient samples were collected and stored in universal transport medium (UTM) or viral transport medium (VTM) and stored at −80C. Samples for luciferase reporter testing were extracted using automatic nucleic acid extraction on the KingFisher Flex Magnetic Particle Processor with 96 Deep Well Head (Thermo Fisher Scientific) using MagMAX™ mirVana™ Total RNA Isolation Kit. For bbLuc, RNA was extracted from 100μL of input volume and eluted into a final volume of 16uL water and stored at −80C. For bbCARMEN, RNA was extracted from 140μL of input volume and eluted into a final volume of 50uL water and stored at −80C.

### Bead preparation and coupling for bbLuc

HiBiT peptides were ordered from Promega as HaloTag protein conjugates (Promega # N3010), or custom-ordered through GenScript Inc. as a peptide with a leading glycine-serine linker (GSSGGSSG-VSGWRLFKKIS) with either an N-terminal azido-lysine modification (for SPAAC) or with an N-terminal maleimide modification (for maleimide-thiol reactions). Biotinylated RNA linkers (supplemental table 1) were attached to the HiBiT peptides using either SPAAC or maleimide-thiol coupling. SPAAC coupling was done at room temperature overnight with a 3:1 ratio of azide-conjugated peptide to DBCO-conjugated oligo; maleimide-thiol coupling was done using a 20:1 ratio of maleimide-conjugated peptide to thiolated oligo left at room temperature for two hours, with an additional overnight incubation at 4C.

For bead coupling, M270 Dynabeads were removed from stock and washed using magnetic separation three times with 1 minute incubations in 1x BW with Tween (5 mM Tris-HCl (pH 7.5), 0.5 mM EDTA, 1 M NaCl, 0.0125% Tween-20). After washing, beads were resuspended in twice the volume of 2x Wash Buffer with Tween (10 mM Tris-HCl (pH 7.5), 1 mM EDTA, 2 M NaCl, 0.025% Tween-20). Beads were then combined in 1X PBS with HaloTag-LgBiT protein (Promega, #CS1967B01) and HaloLigand-Biotin (Promega #G8281) at a 1:1.5 stoichiometric ratio (with HaloTag-LgBiT protein concentration of 80nM unless otherwise noted). Following a 30 minute incubation period on a rotation and another round of bead washing as above, LgBiT-beads were resuspended in Tween buffer (1000uL PBS/.05% Tween 20/0.1% BSA) and HiBiT-beads were resuspended in 1x TEL buffer (0.0125% Tween: 5 mM Tris-HCl (pH 7.5), 0.5 mM EDTA, 10 mM NaCl, 0.0125% Tween-20), followed by storage at 4C.

### Fluorescence and luciferase-based Cas13 assays for bbLuc development

Fluorescence-based reactions were conducted as previously described in (Myhrvold et al. 2018; Barnes et al. 2020; Welch et al. 2022; Arizti-Sanz et al. 2020; Ackerman et al. 2020; Arizti-Sanz et al. 2022) as a point of comparison for luminescent-based reactions and used a polyU FAM reporter. RPA primers and crRNa were designed and selected as described previously ((Myhrvold et al. 2018; Barnes et al. 2020; Welch et al. 2022; Arizti-Sanz et al. 2020; Ackerman et al. 2020; Arizti-Sanz et al. 2022)).

For amplification-free assays, *Leptotrichia wadei Cas13a (LwaCas13a)* protein was first resuspended to 465.7nM in 1X SB (50mM Tris-HCl pH 7.5, 600mM NaCl, 2mM DTT, 5% glycerol). A master mix was created with the following reagents: 1X CB buffer (40mM Tris-HCl (pH 7.5), 1mM DTT), 22.5nM crRNA, 2U/uL RNase Inhibitor Murine (NEB #M0314), 46.6nM *Lwa*Cas13a (GenScript), and 40mM of polyU FAM or 20ug/uL LgBiT and 20ug/uL HiBiT beads. In luciferase reactions, beads were washed in 1x TEL buffer using a process similar to that outlined above (1x TEL buffer: 5 mM Tris-HCl (pH 7.5), 0.5 mM EDTA, 10 mM NaCl, 0.0125% Tween) before being added to the final mix. While the final reaction concentrations of bead bound LgBiT and HiBiT were 80nM and 300nM, respectively, these concentrations were varied in experiments as described above to identify these optimal conditions. Fluorescent or luminescent kinetics were measured at 37C on a Biotek Cytation 5 plate reader using a 384-well Low Flange White Flat Bottom Polystyrene NBS Microplate (3574) or Corning® 384 well microplate (3821), respectively.

For amplification reactions (SHINE), the reaction was done as previously shown in (Myhrvold et al. 2018; Barnes et al. 2020; Welch et al. 2022; Arizti-Sanz et al. 2020; Ackerman et al. 2020; Arizti-Sanz et al. 2022). *Lwa*Cas13a protein was first resuspended to 2250nM in 1X SB (50mM Tris-HCl pH 7.5, 600mM NaCl, 2mM DTT, 5% glycerol). The master mix was created in 1X SHINE buffer (20mM HEPES pH 8.0, 60mM KCl, 5% PEG-8000) included 45nM *Lwa*Cas13a protein, 1U/uL RNase Inhibitor Murine (NEB #M0314), 2 mM of each rNTP (NEB #N0450), 1 U/uL NextGen T7 RNA polymerase, 2U/uL Invitrogen SuperScript IV (SSIV) reverse transcriptase (Thermo Fisher Scientific #18090010), 0.1 U/uL RNase H (NEB #M0297S), 14nM magnesium acetate (Millipore Sigma #63052), 140nM RPA primers, 22.5nM crRNA, and for fluorescence SHINE, 40nM polyU FAM reporter. The reaction was created in reaction units of 107.5uL, with one RPA pellet (TwistDx #TABAS03KIT) per reaction unit.

In the case of luciferase reactions, beads were washed as above in 1x TEL buffer then resuspended in 5mM HEPES buffer, pH 8.0 with 80nM furimazine. To reduce variability caused by reaction viscosity, the reaction master mix was assembled as above and aliquoted to a final reaction tube prior to addition of 20ug/uL LgBiT and HiBiT beads to a final reaction mixture of 19uL. Finally, 1uL target or sample was added to the reaction before measurement for three hours at 37C in a Biotek Cytation 5.

Iterative optimization of the reaction was done via modification of reagent and bead concentration as described in each experiment. Optimal conditions that produced the lowest limit of detection were incorporated into the final protocol as described. In each optimization experiment, the reaction component that was changed is outlined in the results or figures above. The following conditions remained constant across experiments: 45nM *Lwa*Cas13a protein resuspended in 1x SB (such that resuspended protein is at 2.26uM), 1 U/uL murine RNase inhibitor, and 2mM of each rNTP.

For all fluorescent data shown (including curves), fluorescence values were normalized across condition by dividing timepoint data by the mean NTC signal at the first collected timepoint. For luminescence data for amplification-free conditions (including curves) and timepoint curves for SHINE, luminescence values were normalized across condition by dividing timepoint data by the mean NTC signal at the first collected timepoint. For luminesce SHINE data, the larger kinetic complexity precluded the use of a single timepoint to determine a positive/negative call. As such, calls were shown as luminescence ratios, an overall measure of signal across the timepoint curve was determined. This was done by first aligning experimental and NTC condition slopes (as computed between the timepoint nearest to 12 minutes and its subsequent timepoint) by dividing experimental condition by the NTC slope, and next by finding the ratio of the sum of intensities across the experimental and NTC conditions. Patient samples were determined positive with a signal threshold > 1.35.

### Bead preparation and coupling for bbCARMEN

Streptavidin-coated polystyrene beads (Spherotech, no. SVP-200-4) were washed and stored in a binding and washing buffer (2X BW Buffer: 10 mM Tris-HCL pH 7.5, 1 mM EDTA, and 2 M NaCl). To prepare beads for BSA coating, 1mL of beads was washed with 1 mL of 1X BW Buffer three times before being resuspended in 2mL of 2X buffer and 2mL BSA (4mg/mL) (NEB #B9000). Beads were BSA blocked for 3 hours on a rotating stand at room temperature before washing with 1X BW Buffer twice and resuspended in twice the original volume of 2X BW Buffer. BSA blocked beads were stored at 4C until use at a 2.5 ug/uL bead concentration.

crRNA and dye coupling were split into two separate steps. First, 32nM of desired crRNA was mixed with BSA blocked beads in a 1:1 ratio and incubated at room temperature for 15 minutes. After the coupling incubation, crRNA beads were washed with 1X BW Buffer once before resuspending in the original volume of beads with 2X BW Buffer. An equal volume of pre-mixed color-coding dyes (see “color code construction and validation” methods for ratios) were added to the corresponding crRNA bead and incubated at room temperature for 15 minutes. Color-coded crRNA beads were washed six times with 1X BW buffer and then resuspended in 1X TEL buffer (5mM Tris-HCl pH 7.5, 0.5mM EDTA, 10mM NaCl) to have a final bead concentration of 25 ug/uL. crRNA beads were stored at 4C until use and washed twice prior to pooling for each experiment. Equal volumes of beads were pooled together the day of an experiment and incubated in a bbCARMEN Wash Buffer (Cas13 detection master mix without Cas13, 10X Cleavage Buffer, and viral target) for 60 minutes and washed twice with 1X BW Buffer. All washes were accomplished by spinning at 15000 rcf on a tabletop centrifuge and discarding the supernatant. Non-BSA blocked beads required spin times of 3.5 minutes while BSA blocked beads required 1.5 minutes.

### Flow cell design and fabrication for bbCARMEN

Flow cell dimensions were designed in AutoCAD (AutoDesk) and optimized by empirical testing to increase sample size and loading speed. In order to be compatible with existing imaging instruments, the size of a standard microscope slide (25×75mm) was selected. The optimal lane geometry was achieved by maximizing the number of droplets captured in a single lane image field of view. To allow for easy loading, eight 10.5×5.8mm lanes were spaced out on the 75 mm long flow cell with inlet spacing of 9 mm for compatibility with 8-channel multichannel pipettes. Standard size flow cells contain two rows of eight for 16 samples per device. Increasing flow cell dimensions to 50×75 mm enabled 32 lane imaging per device. All flow cells were fabricated with acrylic, a single layer of double sided clear film tape, and hydrophobic treated glass slides. In brief, 12 inch × 12 inch cast acrylic sheets (1⁄4 inch or 1/8 inch, clear) were purchased from Amazon (Small Parts, no. B004N1JLI4) and were cut using an Epilog Fusion M2 laser cutter (60W), producing an acrylic cover with inlets and outlets. Sheets of clear film tape were cut on the laser cutter to provide the geometry of the lanes. Untreated glass slides were treated with Aqualpel from Amazon (Aquapel, no. 2PACK_A) to create a hydrophobic surface. For assembly of the both the 16 and 32 lane flow cells, the clear tape was first adhered to the Aquapel treated glass slide and then to the acrylic cover. Flow cells were stored in plastic bags at room temperature until use.

### Single-step amplification for bbCARMEN

All targets for RVP2.0 were amplified using the QIAGEN OneStep RT-PCR Mix. A total reaction volume of 50 μl was used with some modifications to the manufacturer’s recommended reagent volumes, specifically a 1.25× final concentration of OneStep RT–PCR buffer, 2× more QIAGEN enzyme mix and 20% RNA input. For optimal amplification, final viral primer concentrations varied, with SARS-CoV2, HCoV_NL63, HCoV_OC43, HPIV3, and HMPV primer concentrations at 300nM, HCoV_HKU1 and HRSV at 600nM, FluA and FluB at 480nM, and RNase P at 100nM. The following thermal cycling conditions were used: (1) reverse transcription at 50 °C for 30 min; (2) initial PCR activation at 95 °C for 15 min; and (3) 40 cycles at 94 °C for 30 s, 56 °C for 30 s and 72 °C for 30 s.

### Cas13 detection in bbCARMEN

Detection assays were performed with 45nM purified LwaCas13a, 0.5 ug/uL of pooled crRNA beads, 500nM quenched fluorescent RNA reporter, 1 μl murine 40,000 units/mL RNase inhibitor (New England Biolabs) in nuclease assay buffer (40 mM Tris-HCl, 60 mM NaCl, pH 7.3) with 1mM ATP, 1mM GTP, 1mM UTP, 1mM CTP and 0.6μl T7 polymerase mix (Lucigen).

### Emulsification, loading, and imaging in bbCARMEN flow cells

For emulsification, detection samples (10 uL) were mixed with 2% 008-fluorosurfactant (RAN Biotechnologies) in fluorous oil (3M 7500, 35 μl) in a 96 well plate. Plates were sealed and physically shaken vertically for up to 30 seconds and then spun down for 15 seconds.

For loading, 30 uL of excess oil was removed from each emulsion before 9 uL of droplets were loaded into a 16 or 32 lane bbCARMEN flow cell. The background negative control was computed by analyzing fluorescence signal from droplets containing a scrambled crRNA sequence attached to a color-coded bead. Flow cells were sealed with a PCR film to prevent evaporation of samples.

All bbCARMEN flow cells were imaged on a Nikon TI2 microscope equipped with an automated stage (Ludl Electronics, Bio Precision 3 LM), LED light source (Lumencor, Sola), and camera (Hamamatsu, Orca Flash4.0, C11440, sCMOS) using a 2× objective (Nikon, MRD00025). The following filter cubes were used for imaging: Alexa Fluor 405: Semrock LED-DAPI-A-000; Alexa Fluor 555: Semrock SpGold-B; Alexa Fluor 594: Semrock 3FF03-575/25-25 + FF01-615/24-25; and Alexa Fluor 647: Semrock LF635-B. During imaging, the microscope condenser was tilted back to reduce background fluorescence in the 488 channel. Unless otherwise specified, all flow cells were imaged three times over the course of 60 minutes, with an incubation at 37C between T30 and T60.

### Automated bbCARMEN data analysis

CellProfiler (Stirling et al. 2021) and a custom Jupyter notebook were used to automate image analysis for bbCARMEN (Lamprecht, Sabatini, and Carpenter 2007). First, beads were identified and measured in red, yellow, green, and blue channels using a CellProfiler pipeline. Briefly, images were illumination corrected by subtracting an approximation of image background from each channel. Then, bleedthrough between color channels was computationally compensated for by image subtraction. The corrected images were then masked to exclude the edges of the wells where droplets piled up. Beads were filtered by shape (solidity, eccentricity) to exclude debris and by number of neighbors to exclude beads that were very close to other beads.Beads were also associated with droplets and excluded if a droplet contained multiple beads. The object mask for each accepted bead was expanded 5 pixels and the original bead area subtracted from this to form a ‘donut’ shape in which intensity in the droplet blue channel was measured. For each bead, we calculated normalized intensity measurements for each (red, yellow, green) channel by dividing the mean intensity measurement for each channel by the sum of mean intensities across all 3 channels. Finally, beads were also tracked across images taken at different timepoints using linear assignment problem (LAP) framework (Jaqaman et al. 2008).

CellProfiler measurements were used for bead classification and FAM fluorescence measurement in a separate downstream analysis jupyter notebook. Beads were clustered using normalized red, green, and yellow intensity measurements for each bead using k-means clustering. Results are displayed in a ternary plot showing each bead’s intensity in green, yellow, and red channels and beads are colored by cluster (Supplemental Figure 14). Each cluster was then associated with a virus from the panel by measuring the distance from its measured centroid to the default centroid locations (provided by an external file) and selecting the label of the closest default cluster centroid. In this process, additional QC plots and metrics are also generated and used to assess data quality. Finally, bead donut median blue channel intensities were used to classify samples as positive or negative for each virus in the virus panel using a threshold based on either fold difference from negative control beads (in the same well) and/or exceeding a number of standard deviations above the median intensity of negative control beads. Tracked bead blue channel fluorescence was also plotted over time to observe kinetics of FAM fluorescence.

### SCoV2 RT-qPCR protocol

To detect the presence of SARS-CoV-2 RNA, the extracted RNA samples underwent testing using the CDC’s SARS-CoV-2 RT-qPCR assay (2019-nCoV CDC EUA kit, IDT) targeting the N1 and RP regions. The cycling conditions for the RT-qPCR were as follows: an initial hold at 25 °C for 2 min, followed by reverse transcription at 50 °C for 15 min, polymerase activation at 95 °C for 2 min, and 45 cycles of denaturation at 95 °C for 3s, and annealing/elongation at 55 °C for 30s. The RT-qPCR analysis was performed using a QuantStudio 6 instrument from Applied Biosystems, and the data were analyzed using the Standard Curve module of the Applied Biosystems analysis software.

## Data Availability

All data produced in the present work are contained in the manuscript

## Acknowledgements

We thank the Blainey Lab at the Broad Institute for providing additional lab space for aspects of this work; Tinna-Sólveig Kosoko-Thoroddsen for sharing experimental reagents; and the Sabeti Lab Viral Genomics Group for sharing patient samples. We also thank Lydia Krasilnikova, Brittany Petros, and Elyse Stachler for their thoughtful discussion and feedback on this manuscript. Finally, we note that some elements of schematics shown in this paper were created using www.biorender.com. This work was made possible by funding provided by the Defense Advanced Research Projects Agency (no. D18AC00006), the Centers for Disease Control (no. 75D30122C15113), Howard Hughes Medical Institute, the Flu Lab, and a cohort of donors through the Audacious Project, a collaborative funding initiative housed at TED, including The ELMA Foundation, MacKenzie Scott, the Skoll Foundation, and Open Philanthropy.

