## Supplemental table for "Bead-based approaches to CRISPR diagnostics"

**Table 1**

| **Oligo name** | **Sequence** |
| --- | --- |
| HaloLigand-based linker, 7U | /5Biosg/rUrUrU rUrUrU rU/HaloLigand/ |
| HaloLigand-based linker, 14U | /5Biosg/rUrUrU rUrUrU rUrUrU rUrUrU rUrU/HaloLigand/ |
| HaloLigand-based linker, 21U | /5Biosg/rUrUrU rUrUrU rUrUrU rUrUrU rUrUrU rUrUrU rUrUrU/HaloLigand/ |
| SPAAC linker, 21U, 33 dNTPs | /5Biosg/rUrUrU rUrUrU rUrUrU rUrUrU rUrUrU rUrUrU rUrUrU TTA TTA TTA TTA TTA TTA GGA GGA GCA CGA GGA/3DBCO/ |
| Thiol 21U 7-hexapeg | /5Biosg/rUrU rUrUrU rUrUrU rUrUrU rUrUrU rUrUrU rUrUrU rU/iSp18//iSp18/ /iSp18//iSp18//iSp18/ /iSp18//iSp18//3ThioMC3-D/ |
| Thiol 2-hexapeg 14 U | /5Biosg//iSp18//iSp18//rUrUrU rUrUrU rUrUrU rUrUrU rUrU/3ThioMC3-D/ |
| Thiol 3-hexapeg 10 U | /5Biosg//iSp18//iSp18//iSp18//rUrUrU rUrUrU rUrUrU rU/3ThioMC3-D/ |
| Thiol 4-hexapeg 6U | /5Biosg//iSp18//iSp18//iSp18//iSp18//rUrUrU rUrUrU/3ThioMC3-D/ |
| SPAAC 2-hexapeg 14 U | /5Biosg//iSp18//iSp18/rUrUrU rUrUrU rUrUrU rUrUrU rUrU/3DBCON/ |
| SPAAC 3-hexapeg 10 U | /5Biosg//iSp18//iSp18//iSp18/rUrUrU rUrUrU rUrUrU rU/3DBCON/ |
| SPAAC 4-hexapeg 6U | /5Biosg//iSp18//iSp18//iSp18//iSp18/rUrUrU rUrUrU/3DBCON/ |
| SPAAC 4-hexapeg 14 U | /5Biosg//iSp18//iSp18//iSp18//iSp18/rUrUrU rUrUrU rUrUrU rUrUrU rUrU/3DBCON/ |
| Thiol 2-hexapeg 21 U | /5Biosg//iSp18//iSp18//rUrUrU rUrUrU rUrUrU rUrUrU rUrUrU rUrUrU rUrUrU/3ThioMC3-D/ |
| Thiol 4-hexapeg 21 U | /5Biosg//iSp18//iSp18//iSp18//iSp18//rUrUrU rUrUrU rUrUrU rUrUrU rUrUrU rUrUrU rUrUrU/3ThioMC3-D/ |
| Spike 69/70 gBlock | gaaatTAATACGACTCACTATAggTGACAAAGTTTTCAGATCCTCAGTTTTACATTCAACTCAGGACTTGTTCTTACCTTTCTTTTCCAATGTTACTTGGTTCCATGCTATACATGTCTCTGGGACCAATGGTACTAAGAGGTTTGATAACCCTGTCCTACCATTTAATGATGGTGTTTATTTTGCTTCCACTGAGAAGTCTAACATAATAAGAGGCTGGATTTTTGGTACTACTTTAGATTCGAAGACCCAGTCCCTACTTATTGTTAATAACGCTACTAATGTTGTTATTAAAGTCTGTGAATTTC |
| Spike RPA primer forward | gaaatTAATACGACTCACTATAgggCAACTCAGGACTTGTTCTTACCTTTCTTTTCC |
| Spike RPA primer reverse | AAGCAAAATAAACACCATCATTAAAT |
| Spike crRNA 69/70 | rGrArU rUrUrA rGrArC rUrArC rCrCrC rArArA rArArC rGrArA rGrGrG rGrArC rUrArA rArArC rArCrA rGrGrG rUrUrA rUrCrA rArArC rCrUrC rUrUrA rGrUrA rCrCrA rU |
| FAM_7U_Reprter | /56-FAM/rUrUrUrUrUrUrU/3IABkFQ/ |
| SARS-CoV-2 crRNA | GAUUUAGACUACCCCAAAAACGAAGGGGACUAAAACCUAAAACUAUUCACUUCAAUAGUCUGAA/3Bio/ |
| HCoV-HKU1 crRNA | GAUUUAGACUACCCCAAAAACGAAGGGGACUAAAACAAUAUGAUUACCAUUACCACAAAAAUUA/3Bio/ |
| HCoV-NL63 crRNA | GAUUUAGACUACCCCAAAAACGAAGGGGACUAAAACUUAAUAGUUUCAGCCGCAAAGAGUCUAA/3Bio/ |
| HCoV-OC43 (BetaCoV) crRNA | GAUUUAGACUACCCCAAAAACGAAGGGGACUAAAACUGUUGUAACGCCCUUAUAAUAGACCUUA/3Bio/ |
| HPIV3 crRNA | GAUUUAGACUACCCCAAAAACGAAGGGGACUAAAACGUCGCAUUUUCCCCUCAAUAGAGUCCUU/3Bio/ |
| FluA crRNA | GAUUUAGACUACCCCAAAAACGAAGGGGACUAAAACAAAAAGCUUGUGAAUUCAAAUGUCCCUG/3Bio/ |
| FluB crRNA | GAUUUAGACUACCCCAAAAACGAAGGGGACUAAAACACUAAACAGAUCAGGACAAGGUAUUUGG/3Bio/ |
| HMPV crRNA | GAUUUAGACUACCCCAAAAACGAAGGGGACUAAAACGUCGCAAAAGACAUGGUCUCCUCUUGUU/3Bio/ |
| HRSV crRNA | GAUUUAGACUACCCCAAAAACGAAGGGGACUAAAACGUCUUUUUCUAGGACAUUGUAUUGAACA/3Bio/ |
| RNaseP crRNA | GAUUUAGACUACCCCAAAAACGAAGGGGACUAAAACUCCGAGUCAGUGGCUCCCGUGUGUCGGU/3Bio/ |
| Scrambled 1 crRNA | GAUUUAGACUACCCCAAAAACGAAGGGGACUAAAACACGUCUAAUACGAUACAUCAUUACAUAU/3Bio/ |
| Scrambled 2 crRNA | GAUUUAGACUACCCCAAAAACGAAGGGGACUAAAACGUGCGCCGUUGGCUCGUGUAGCAGUUCC/3Bio/ |
| SARS-CoV-2 forward primer | gaaatTAATACGACTCACTATAgggCAATTAGAGATGGAACTTACACC |
| HCoV-HKU1 forward primer | gaaatTAATACGACTCACTATAgggGTGTGTTAAAAGTCAATCTCCTCG |
| HCoV-NL63 forward primer | gaaatTAATACGACTCACTATAgggACTTGCTAATGATGTTAAAGATACAC |
| HCoV-OC43 (BetaCoV) forward primer | gaaatTAATACGACTCACTATAgggGCTAAGAATGAGAGTAGTTCATTG |
| HPIV3 forward primer | gaaatTAATACGACTCACTATAgggTGATCTCAATGAAATTAGAAAGATGG |
| FluA forward primer | gaaatTAATACGACTCACTATAgggGAGCAAAAAGAAGTCCTATATAAATAAG |
| FluB forward primer | gaaatTAATACGACTCACTATAgggCAAGCAAAACAAAAAGACTAAAGGC |
| HMPV forward primer | gaaatTAATACGACTCACTATAgggACCCAAATGAGAAAGACTGTG |
| HRSV forward primer | gaaatTAATACGACTCACTATAgggCTTCACGAAGGCTCCACATA |
| RNaseP forward primer | gaaatTAATACGACTCACTATAgggTTGATGAGCTGGAGCCA |
| SARS-CoV-2 reverse primer | CTTTTTAGCTTCTTCCACAATGTC |
| HCoV-HKU1 reverse primer | AACCATAAGGAGCATTTTGAAC |
| HCoV-NL63 reverse primer | GACTTAACACTCTCTTCTTTAGCT |
| HCoV-OC43 (BetaCoV) reverse primer | ATTTACAGCACTAGAACTTTCATG |
| HPIV3 reverse primer | CTGATATCTCGCTTGGAACATCTGCAG |
| FluA reverse primer | AATTAGCCACAAATCCATAGCG |
| FluB reverse primer | TGTTTCTTCATTATATCTTTCTAATGGTAT |
| HMPV reverse primer | GCAACATTAATTCCTGCTGCT |
| HRSV reverse primer | CCCATATTGTTAGTGATGCAGG |
| RNaseP reverse primer | ATGTGGATGGCTGAGTTGTT |
| SARS-CoV-2 gBlock | GCACCCATATTGTTAGTG |
| HCoV-HKU1 gBlock | gaaatTAATACGACTCACTATAgggATGCTCTTCTTTCTATTCAGAATGGTTTTAGTGCTACCAACTCTGCACTTGCTAAAATACAAAGTGTTGTTAATTCTAATGCTCAAGCACTTAATAGTTTGTTACAGCAATTATTTAATAAATTTGGTGCAATTAGTTCTTCTTTACAAGAAATTTTATCTCGTCTCGATGCTTTAGAGGCTCAGGTTCAGATTGATAGGCTTATTAATGGTCGTTTAACTGCTTTAAATGCTTATGTCTCTCAACAGCTTAGTGATATTTCTCTTGTAAAATTTGGTGCTGCTTTAGCTATGGAGAAGGTTAATGAGTGTGTTAAAAGTCAATCTCCTCGTATTAATTTTTGTGGTAATGGTAATCATATTTTGTCATTAGTTCAAAATGCTCCTTATGGTTTGTTGTTTATGCATTTTAGTTATAAACCTATTTCTTTTAAAACTGTTTTAGTAAGTCCTGGTTTGTGTATATCAGGTGATGTAGGTATTGCACCTAAACAAGGGTAT |
| HCoV-NL63 gBlock | gaaatTAATACGACTCACTATAgggACGTTATGTGTCTTTAGCTATTGATGCATACCCTCTTTCAAAACACCCTAATTCTGAATATCGTAAGGTTTTTTACGTATTACTTGATTGGGTTAAGCATCTTAACAAAAATTTGAATGAGGGTGTTCTTGAATCTTTTTCTGTTACACTTCTTGATAATCAAGAAGATAAGTTTTGGTGTGAAGATTTTTATGCTAGTATGTATGAAAATTCTACAATATTGCAAGCTGCTGGTTTATGTGTTGTTTGTGGTTCACAAACTGTACTTCGTTGTGGTGATTGTCTGCGTAAGCCTATGTTGTGCACTAAATGCGCATATGATCATGTATTTGGTACCGACCACAAGTTTATTTTGGCTATAACACCGTATGTATGTAATGCATCAGGTTGTGGTGTTAGTGATGTCAAAAAATTGTATCTTGGTGGTTTGAATTACTATTGTACAAATCATAAACCACAGTTGTCTTTTCCATTATGTTCAGCTGGTAATATATTTGGTTTATATAAAAATTCAGCAACTGGTTCCTTAGATGTTGAAGTTTTTAATAGGCTTGCAACGTCTGATTGGACTGATGTTAGGGACTATAAACTTGCTAATGATGTTAAAGATACACTTAGACTCTTTGCGGCTGAAACTATTAAAGCTAAAGAAGAGAGTGTTAAGTCTTCTTATGCTTTTG |
| HCoV-OC43 (BetaCoV) gBlock | gaaatTAATACGACTCACTATAgggGTTGTAGATGAAGTTAGCATGCTTACCAATTATGAGCTTTCTGTTATTAATGCTCGTATTCGTGCTAAGCATTATGTTTATATTGGTGATCCTGCTCAATTGCCAGCACCACGTGTGTTATTGAGCAAGGGTACACTTGAACCTAAATATTTTAACACTGTTACTAAGCTCATGTGTTGCTTAGGGCCAGACATTTTTCTTGGTACATGTTATAGATGTCCTAAGGAAATTGTTGATACAGTGTCCGCCTTGGTTTATGAAAATAAGCTTAAGGCTAAGAATGAGAGTAGTTCATTGTGTTTTAAGGTCTATTATAAGGGCGTTACAACACATGAAAGTTCTAGTGCTGTAAATATGCAGCAGATTTATTTGATTAATAAGTTTTTGAAGGCTAACCCTTTGTGGCATAAAGCTGTTTTTATTAGCCCATATAATAGTCAGAACTTTGCAGCTAAGCGTGTTTTGGGTTTACAAACCCAAACCGTGGATTCTGCTCAAGG |
| HPIV3 gBlock | gaaatTAATACGACTCACTATAgggACCATCTGTCAACCAGAAATCAAACCAACAGAAACAAGTGAAAAAGATAGTGGATCAACTGACAAAAATAGACAGTCTGGGTCATCACACGAATGTACAACAGAAGCAAAAGATAGAAATATTGATCAGGAAACTGTACAGAGAGGACCTGGGAGAAGAGGCAGCTCAGATAGTAGAGCTGAGACTGTGGTCTCTGGAGGAATCTCCAGAAGCATCACAGATTCTAAAAATGGAACCCAAAACACGGAGAATATTGATCTCAATGAAATTAGAAAGATGGATAAGGACTCTATTGAGGGGAAAATGCGACAATCTGCAGATGTTCCAAGCGAGATATCAGGAAGTGATGGCATATTTACAACAGAACAAAGTAGAAACAGTGATCATGGAAGAAGCTTGGAATCTATCGGTACACCTGATACAAGATCAATAAGTGTTGTTACTGCTGCAACACCAGATGATGAAGAAGAAATACTAATGAGAAATAGTAGGATGAAGAA |
| FluA gBlock | gaaatTAATACGACTCACTATAgggTGAATCAACAAGGAAGAAAATTGAGAAGATAAGGCCTCTTTTAATGGATGGCACAGCATCACTGAGTCCTGGGATGATGATGGGCATGTTCAACATGCTAAGTACAGTCTTGGGAGTCTCGATACTGAATCTTGGACAAAAGAAATACACCAAGACAACATACTGGTGGGATGGGCTCCAATCATCCGACGATTTTGCTCTCATAGTGAATGCACCAAACCATGAAGGAATACAAGCAGGAGTGGACAGATTCTACAGGACCTGCAAATTAGTGGGAATCAACATGAGCAAAAAGAAGTCCTATATAAATAAGACAGGGACATTTGAATTCACAAGCTTTTTTTATCGCTATGGATTTGTGGCTAATTTTAGCATGGAGCTACCCAGCTTTGGAGTGTCTGGAGTAAATGAATCAGCTGACATGAGTATTGGAGTAACAGTGATAAAGAACAACATGATAAACAATGACCTTGGACCTGCAACGGCTCAGATGGCTCTTC |
| FluB gBlock | gaaatTAATACGACTCACTATAgggCAAGCAAAACAAAAAGACTAAAGGCCCAAATACCTTGTCCTGATCTGTTTAGTATACCATTAGAAAGATATAATGAAGAAACAAGGGCAAAATTGAAGAAGCTAAAACCATTCTTCAATGAAGAAGGAACTGCATCTTTGTCACCTGGGATGATGATGGGAATGTTTAATATGCTATCTACCGTGTTGGGAGTAGCTGCACTAGGTATCAAGAACATTGGAAACAAAGAATACCTATGGGATGGACTGCAATCTTCTGATGATTTTGCTCTATTTGTTAATGCAAAGGATGAAGAAACATGTATGGAAGGAATAAACGACTTTTACCGAACATGTAAATTATTGGGAATAAACATGAGCAAAAAGAAAAGTTACTGTAATGAGACTGGAATGTTTGAATTTACAAGCATGTTCTACAGAGATGGATTTGTATCTAATTTTGCAATGGAACTCCCTTCGTTTGGGGTTGCTGGAGTAAATGAATCAGCAGATATGGCAATA |
| HMPV gBlock | gaaatTAATACGACTCACTATAgggGAGAAGACCAAGGGTGGTATTGTCAGAATGCAGGGTCAACTGTTTACTACCCAAATGAGAAAGACTGTGAAACAAGAGGAGACCATGTCTTTTGCGACACAGCAGCAGGAATTAATGTTGCTGAGCAATCAAAGGAGTGCAACATCAACATATCCACTACAAATTACCCATGCAAAGTCAGCACAGGAAGACATCCTATCA |
| HRSV gBlock | gaaatTAATACGACTCACTATAgggGGGGCAAATATGGAAACATACGTGAACAAACTTCACGAAGGCTCCACATACACAGCTGCTGTTCAATACAATGTCCTAGAAAAAGACGATGATCCTGCATCACTTACAATATGGGTGCCCATGTTCCAATCATCCATGCCAGCAGATTTACTTATAAAAGAACTAGCTAATGTCAACATACTAGTGAAACAAATATCCACA |
| RNaseP gBlock | gaaatTAATACGACTCACTATAgggTCCTTGCAGGTGGCTGCCAATACCTCCACCGTGGAGCTTGTTGATGAGCTGGAGCCAGAGACCGACACACGGGAGCCACTGACTCGGATCCGCAACAACTCAGCCATCCACATCCGAGTCTTCAGGGTCACACCCAAGTAATTGAAAAGACACTCCTCCACTTATCCCCTCCGTGATATGGCTCTTCGCATGCTGAGTACTGGACCTCGGACCAGAGCCATGTAAGAAAAGGCCTGTTCCCTGGAAGCCCAAAGGACTCTGCATTGAGGGTGGGGGTAATTGTCTCTTGGTGGCCCAGTTAGTGGGCCTTCCTGA |
