## Supplementary Figures for "Bead-based approaches to CRISPR diagnostics"

#### Supplementary Figure 1: Fluorescence vs split Luminescent systems

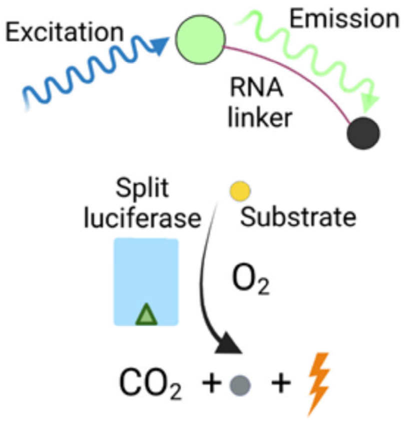


**Supplementary Figure 1:** Schematic of quenched fluorescence vs split luciferase-based reporter systems.

### Supplementary Figure 2: Optimizing LgBiT Concentration

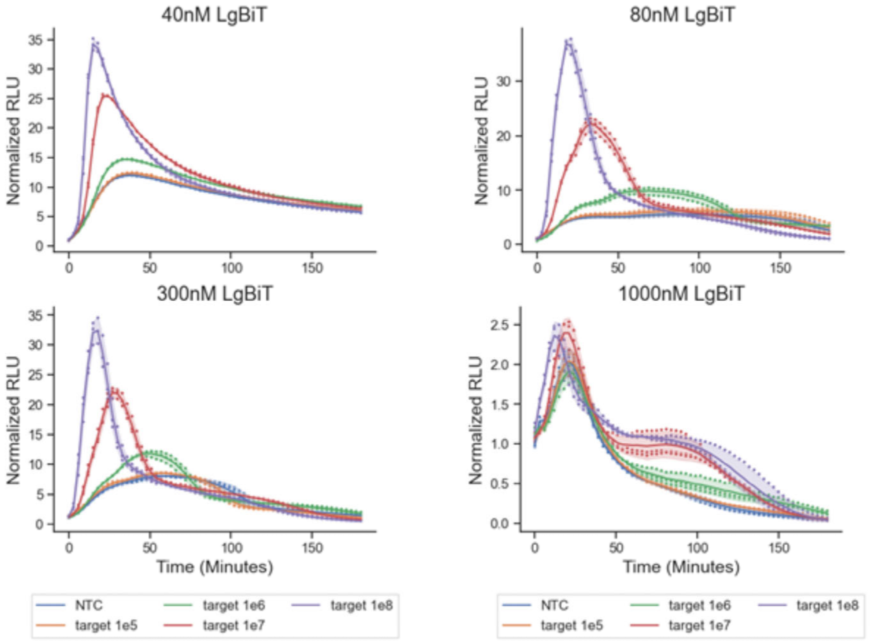


**Supplementary Figure 2: Optimizing LgBiT Concentration.** Detection-only luminescent reaction with different concentrations of LgBiT (40 nM, 80 nM, 300 nM, and 1000 nM) and 300 nM HiBiT after 3h on varied synthetic RNA target; NTC, no target control.

80nM and 40nM LgBiT concentrations showed similar detection efficiencies, with 1000nM LgBiT-np showing heavily reduced detection, likely due to saturation of LgBiT-np and HiBiT-np bead-bead interactions. 80nM LgBiT was ultimately chosen as it demonstrated a higher SNR of any of the conditions tested.

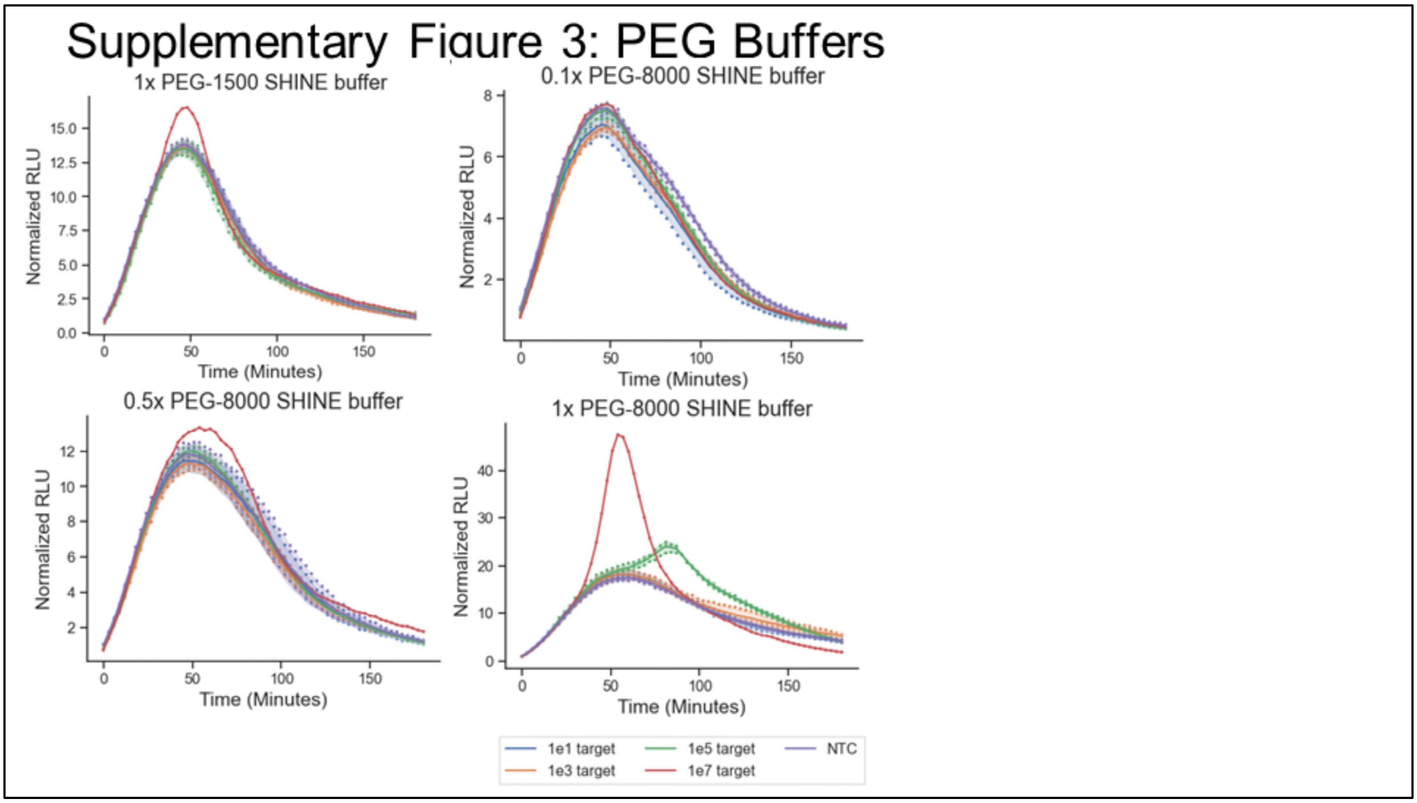


**Supplementary Figure 3: Optimizing PEG Buffer Concentration.** Luminescent SHINE was performed with varying makeup of PEG buffer, specifically 5% PEG-8000, 2.5% PEG-8000, 0.5% PEG-8000, and 5% PEG-1500 on synthetic RNA target; NTC, no target control. Optimal performance was observed with 5% PEG-8000, with other concentrations severely limiting performance.

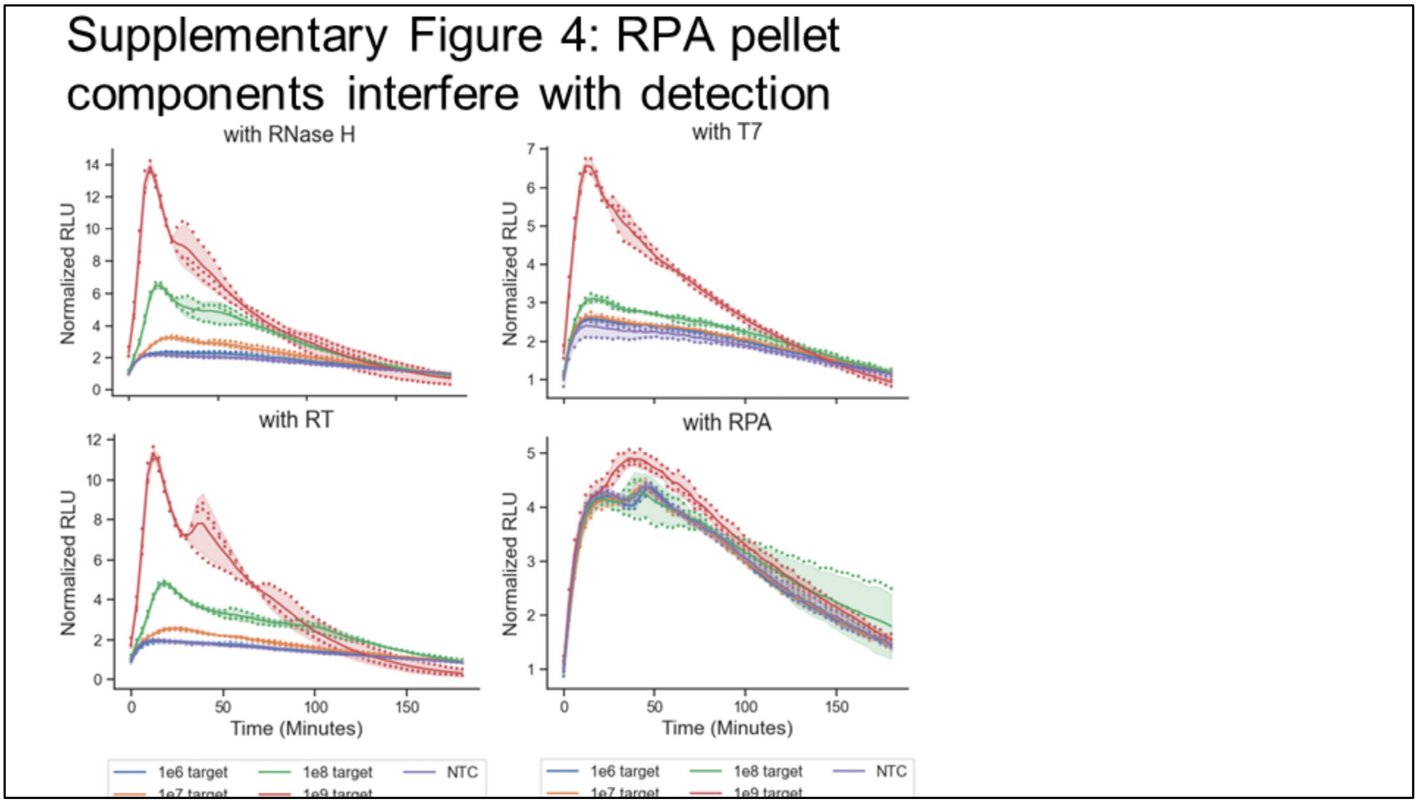


**Supplementary Figure 4: Components of RPA pellets interfere with detection.** Amplification-free reactions in SHINE buffer, with RPA pellets, RNase H, reverse transcriptase, and T7 RNA polymerase spiked in separately in different conditions. RNase H, reverse transcriptase, T7 RNA polymerase additions show little relative inhibition to detection in SHINE buffer. In contrast, addition of RPA pellets significantly degrades detection-only performance. Experiments done on varied synthetic RNA target; NTC, no target control.

#### Supplementary Figure 5: SSB and recombinases in RPA interfere with detection

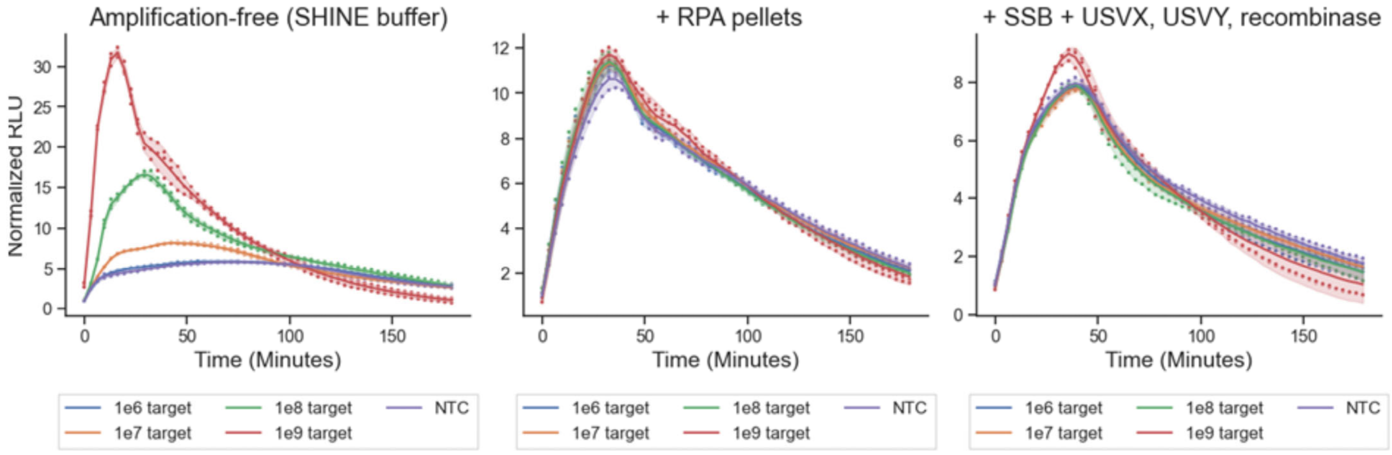


**Supplementary Figure 5: RPA pellet components (SSB and recombinases) interfere in detection.** Luminescent amplification-free assays, with optimized CB buffer substituted with SHINE buffer. Differing conditions with RPA pellets spiked in, and with constituent enzymes of RPA (SSB, USVX, USVY) added in RPA concentrations.

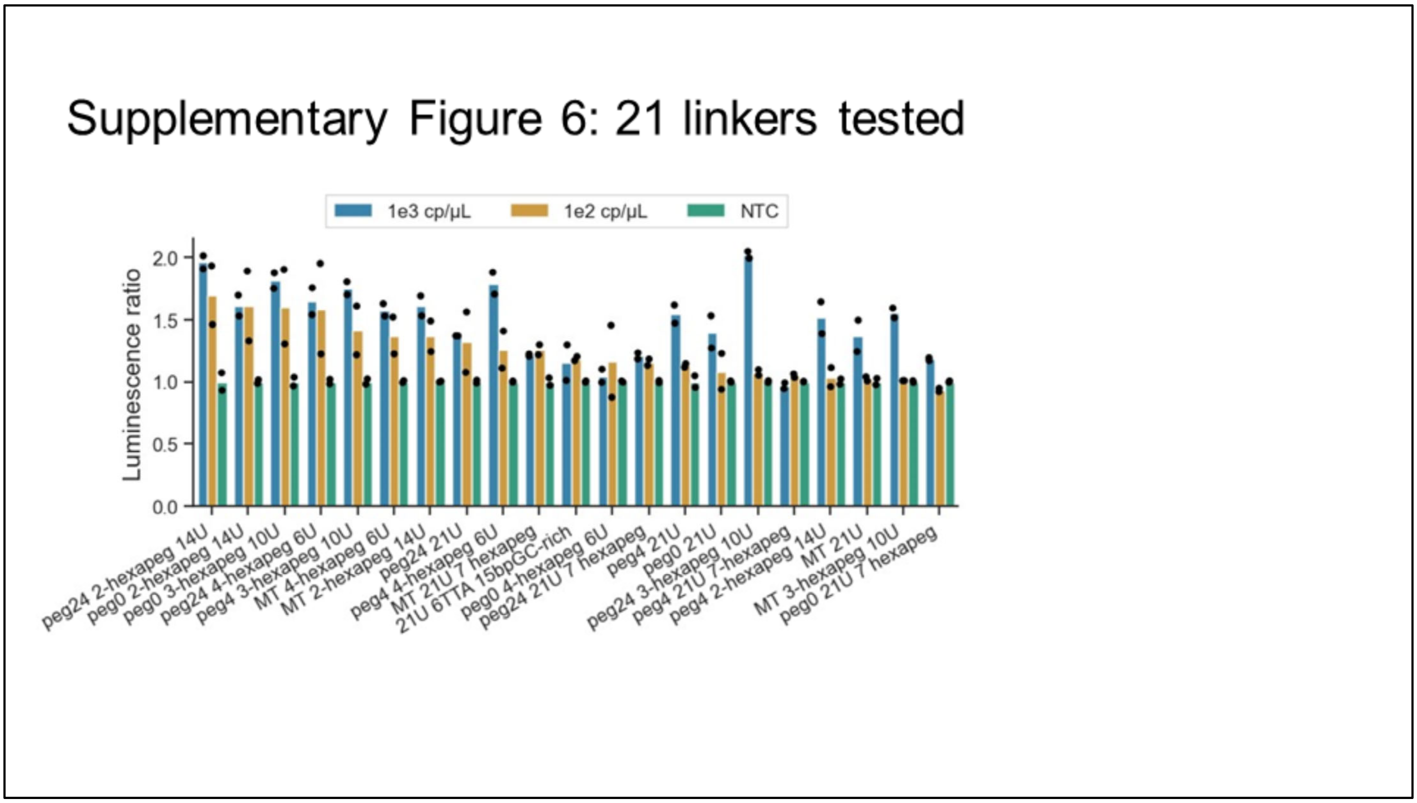


**Supplementary Figure 6: Detection using 21 different RNA linkers.** We tested several different designs of oligos with different types of conjugation (SPAAC, MT, DBCO-PEG-MT) and 8 types of oligos. Five different base oligos were used (xo1, R50, R51, R52, R53). Three different conjugation methods were also used (Maleimide-Thiol and Maleiemide-Thiol-PEG-DBCO). The bars are ordered as descending ratios of 1e2 target to NTC signal.

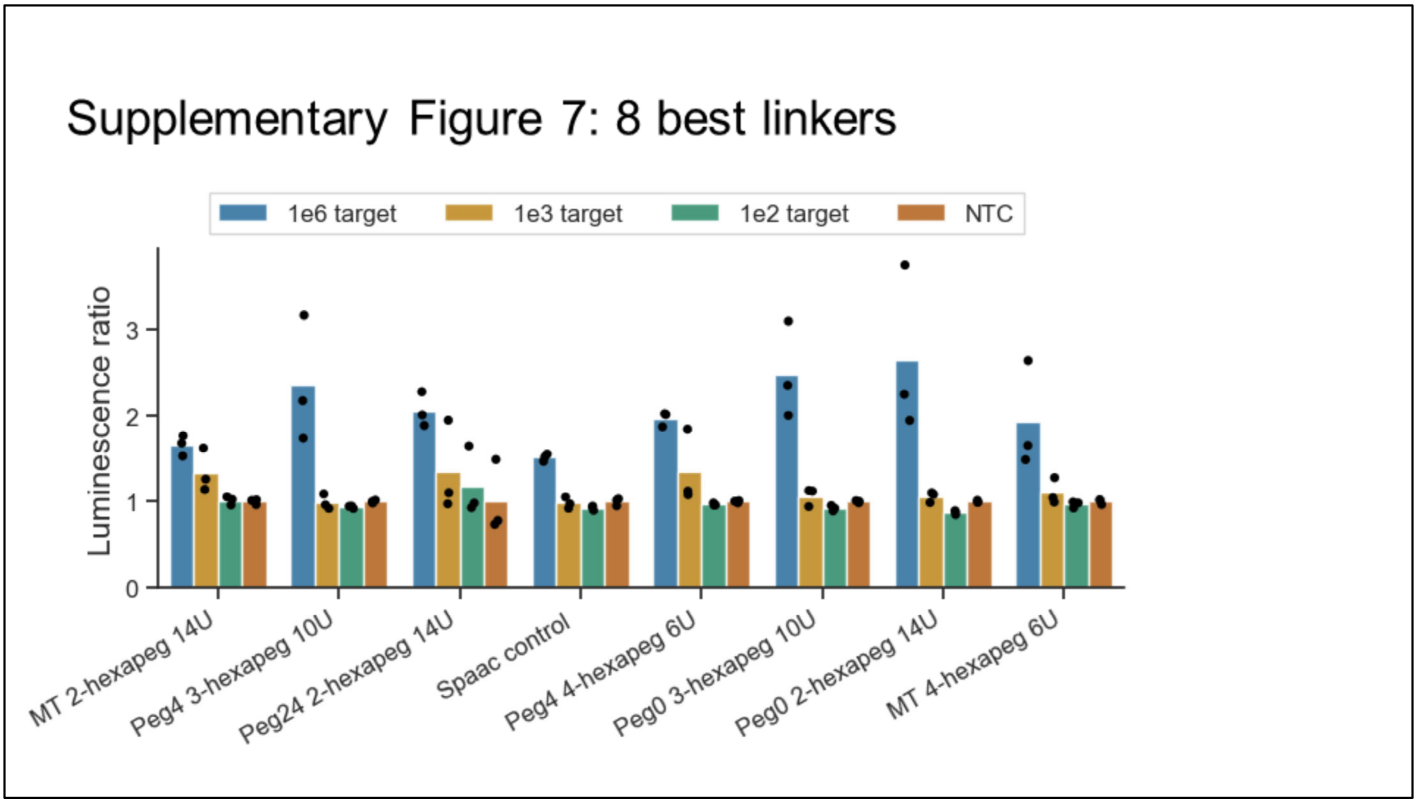


**Supplementary Figure 7: Detection using 8 best RNA linkers.** We tested several different designs of oligos with different types of conjugation (SPAAC, MT, DBCO-PEG-MT) and 8 types of oligos.

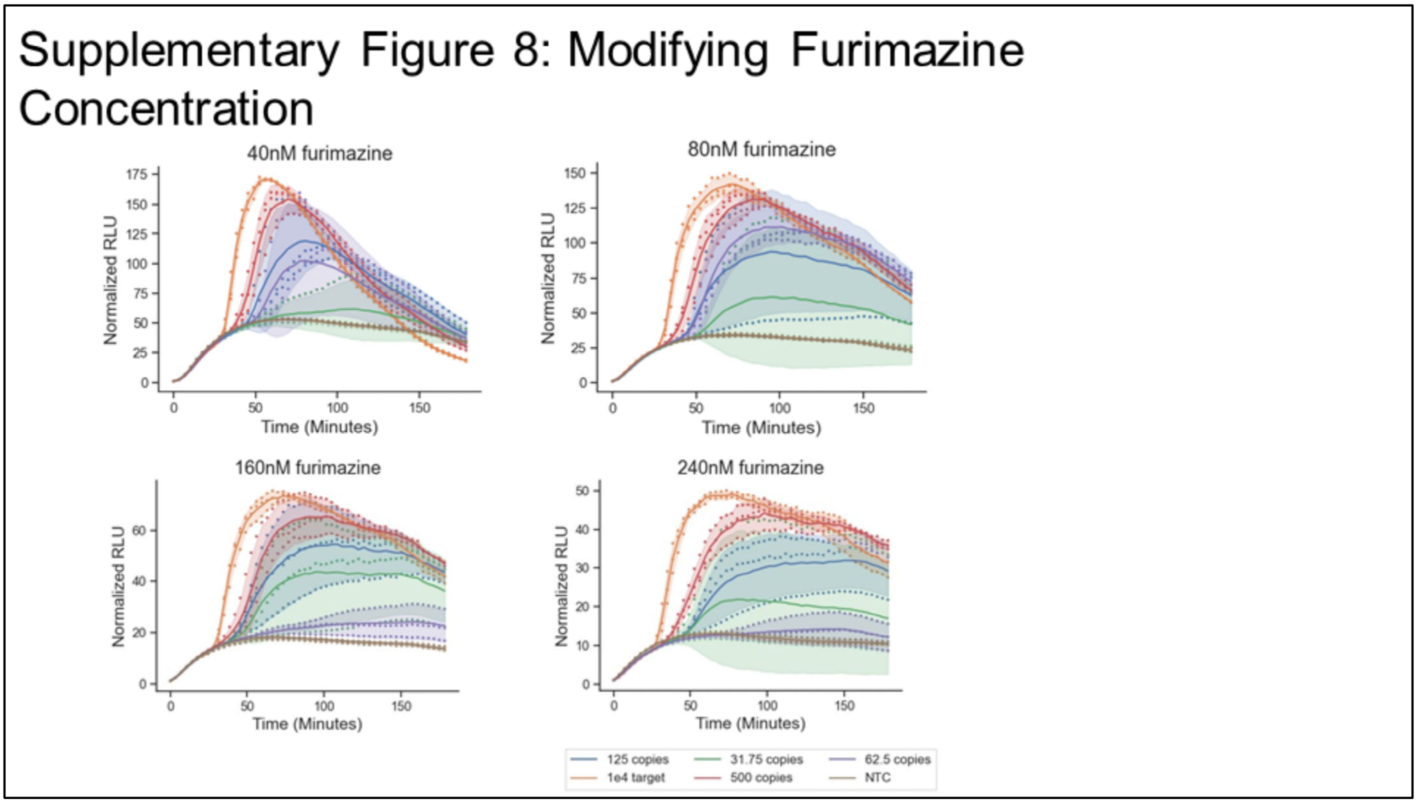


**Supplementary Figure 8: Optimizing furimazine concentration.** Luminescent SHINE assay showing different concentrations of furimazine (40nM, 80nM, 160nM, 240nM).

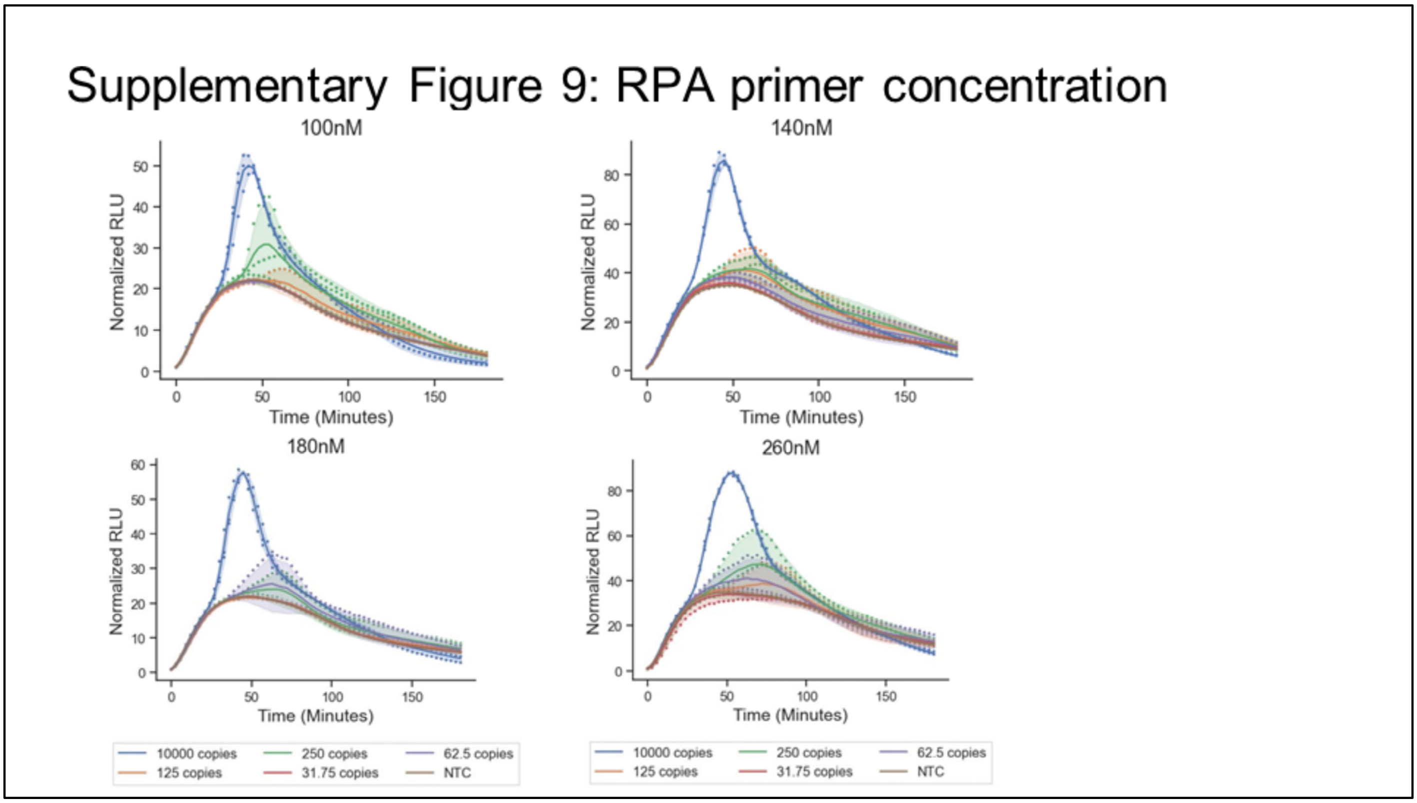


**Supplementary Figure 9: Optimizing RPA primer concentration.** Luminescent SHINE assay showing different concentrations of RPA primers (100nM, 140nM, 180nM, 260nM).

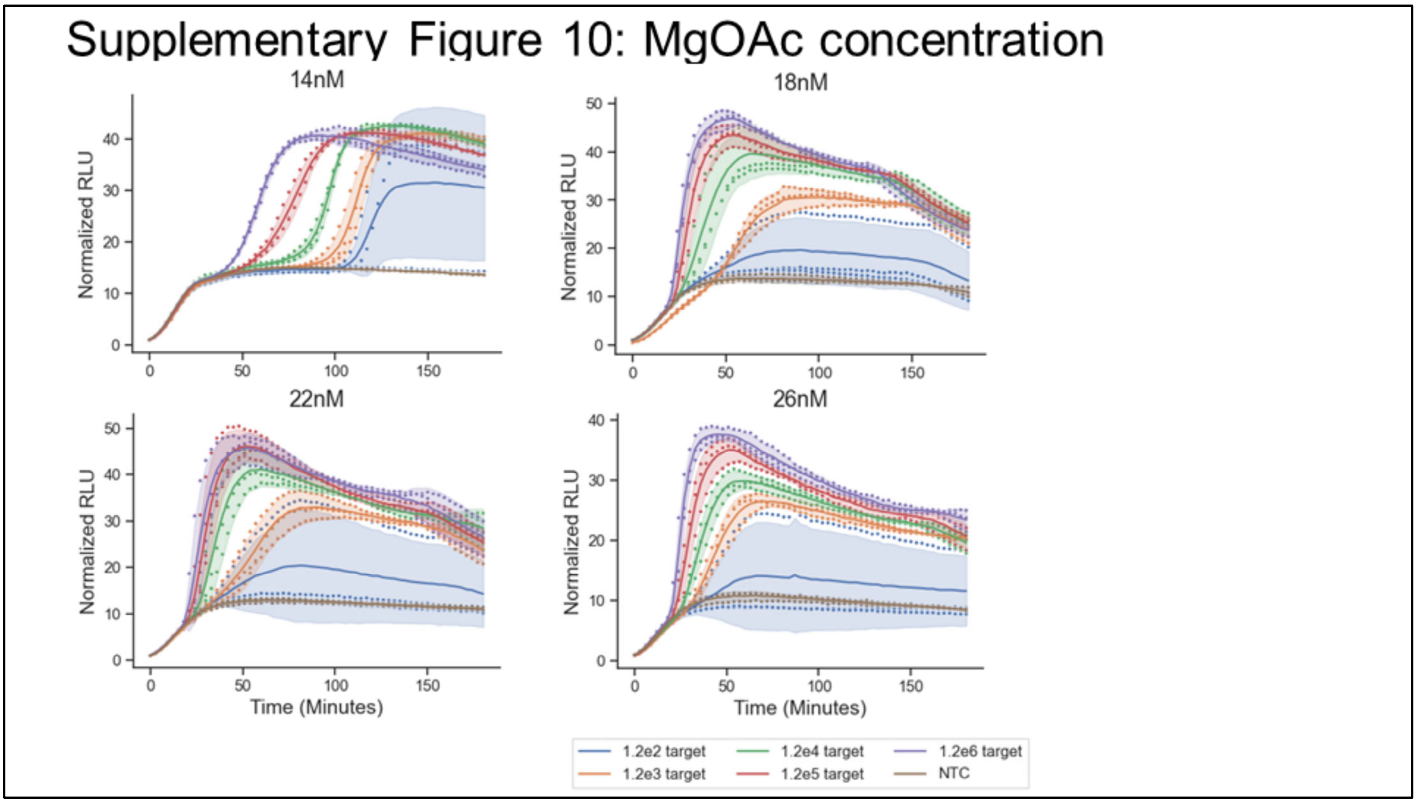


**Supplementary Figure 10: Optimizing magnesium acetate concentration**. We tested different concentrations of magnesium acetate. We found that 14nM of MgOAc has the best sensitivity, however 18 nM has better speed (faster detection prior to 40 minutes).

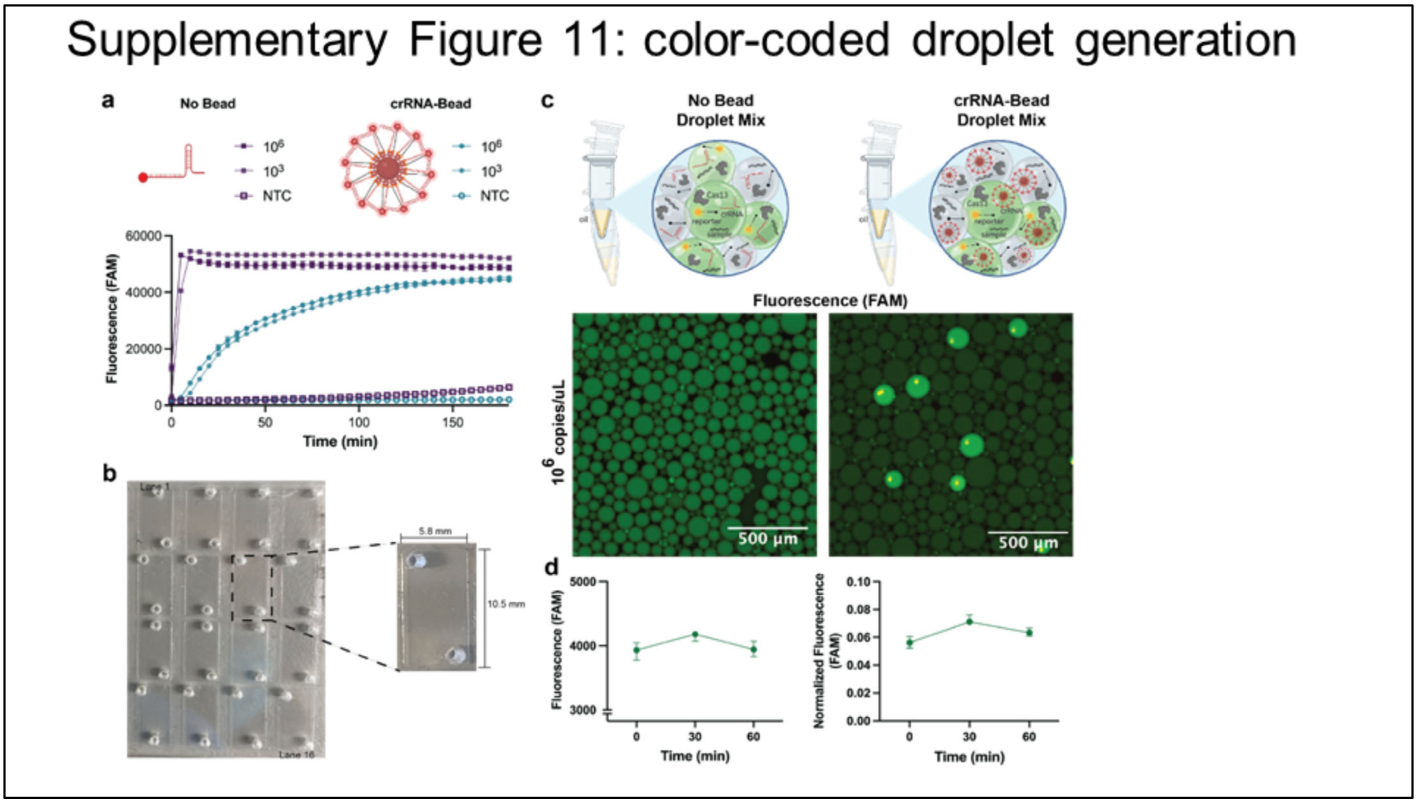


**Supplementary Figure 11. Equipement-free droplet generation with color-coded crRNA beads for Cas13-based detection. a**, Kinetics of SARS-CoV-2 at $10^6$ and $10^3$ copies/uL with SARS-CoV-2 crRNA either bound or not bound to a biotinylated bead. Fluorescence measured on the Cytation 5 plate reader. **b**, Image of flow cell with 5.8 x 10.5 mm lane dimensions that can be loaded using a multichannel pipette. Flow cells fabricated with 16 lanes, 25 x 75 mm, or 32 lanes, 50 x 75 mm. Shown as 16 lanes in **b**. **c**, Fluorescent images of droplets in the absence of beads (left) or presence of crRNA-beads (right) SARS-CoV-2 at $10^6$ copies/uL. **d**, Fluorescence kinetics of SARS-CoV-2 crRNA in droplets with or without bead attachment and compared to signal from a no droplet control.

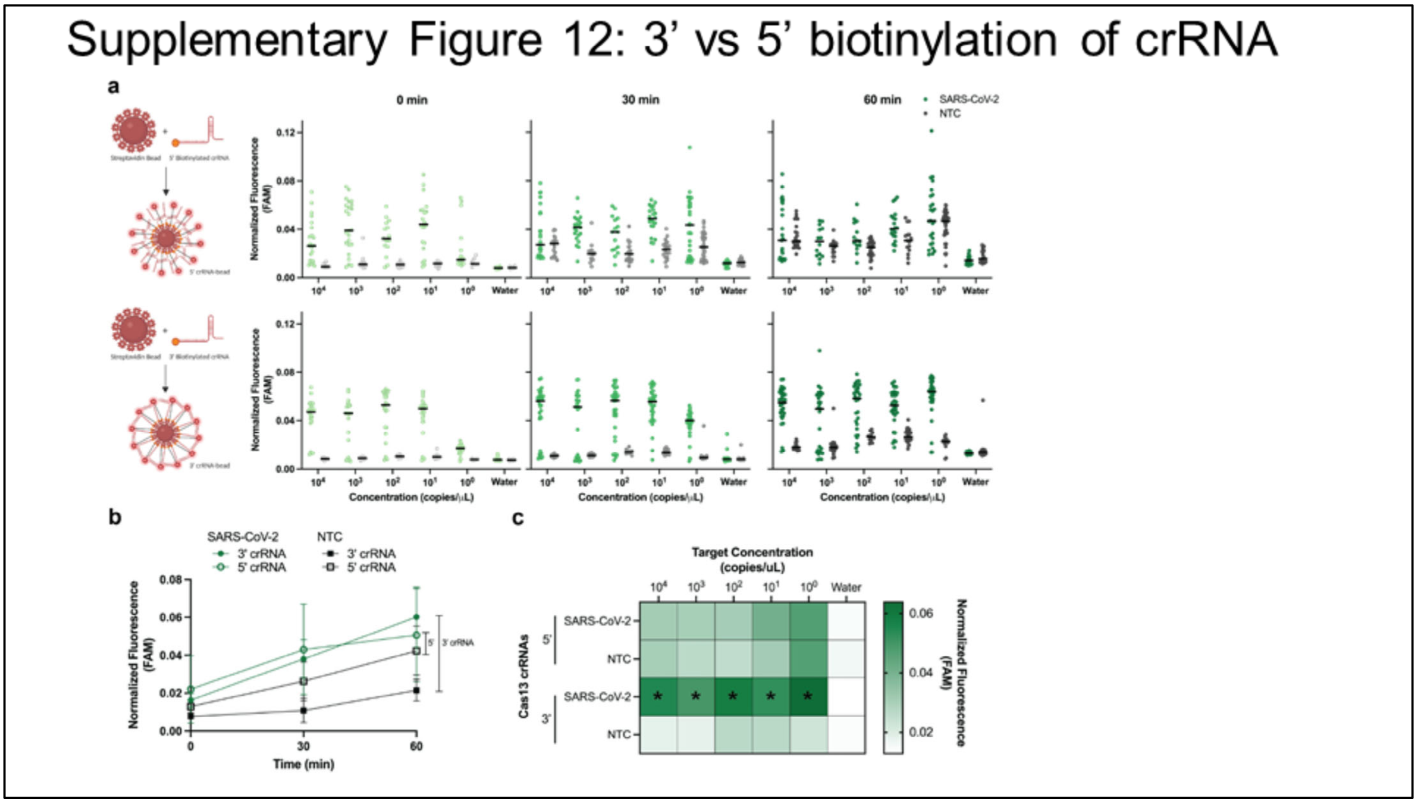


**Supplementary Figure 12. Background signal reduction with 3' biotinylated crRNA-bead pools compared to 5' pools. a**, Fluorescence across amplified SARS-CoV-2 dilution series from $10^4$-$10^0$ copies/uL at 0, 30, and 60 min post-reaction initiation. Top: 5' biotinylated crRNA-bead pools; Bottom: 3' biotinylated crRNA-bead pools; Green: SARS-CoV-2; Gray: NTC. Bar at median fluorescence. **b**, Fluorescence kinetics of SARS-CoV-2 at $10^0$ copies/uL from 3' and 5' biotinylated crRNA-bead pools. Green: SARS-CoV-2; Black: NTC; Closed points: 3' biotinylated crRNA; Open points: 5' biotinylated crRNA. **c**, Heatmap of median SARS-CoV-2 and NTC fluorescence at 60 min post-reaction initiation from **a**. Asterisk (*) represents positive signal detected above threshold.

### Supplementary Figure 13: Dual SCoV2 & Rnase P

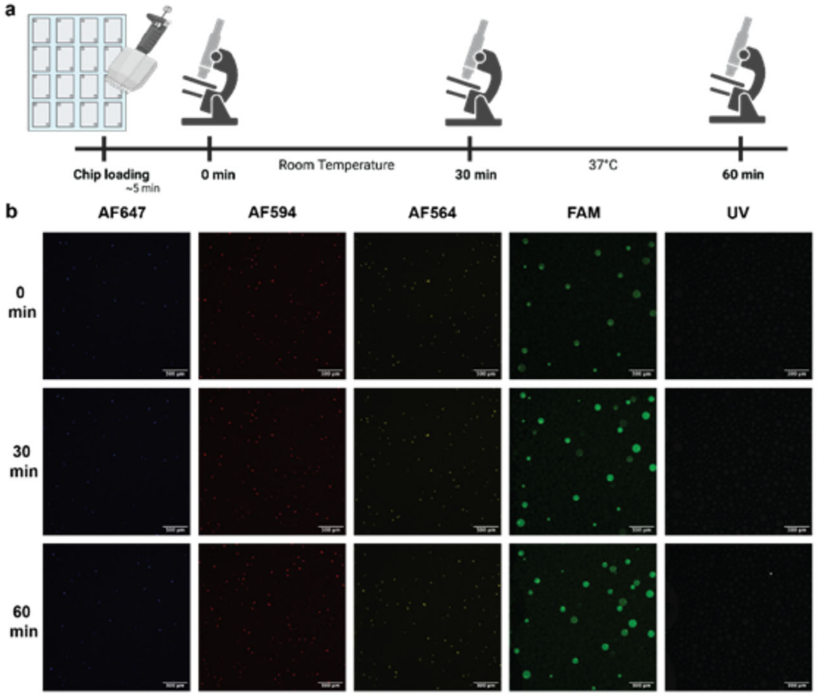


**Supplementary Figure 13. Dual SARS-CoV-2 and RNase P fluorescence imaging over time. a**, Schematic of bbCARMEN flow cell imaging up to 60 min post-reaction initiation. **b**, Fluorescent images at 0, 30, and 60 min post-reaction in 4 different fluorescent channels. Synthetic SARS-CoV-2 RNA at $10^6$ copies/uL was spiked into RNase P. Blue: AF647; Red: AF594; Yellow: AF564; Green: FAM. AF647: Semrock LF635-B; AF594: Semrock 3FF03-575/25-25 and FF01-615/24-25; AF546: Semrock SpGold-B; FAM: Semrock GFP-1828A. Scale bar: 500 um.

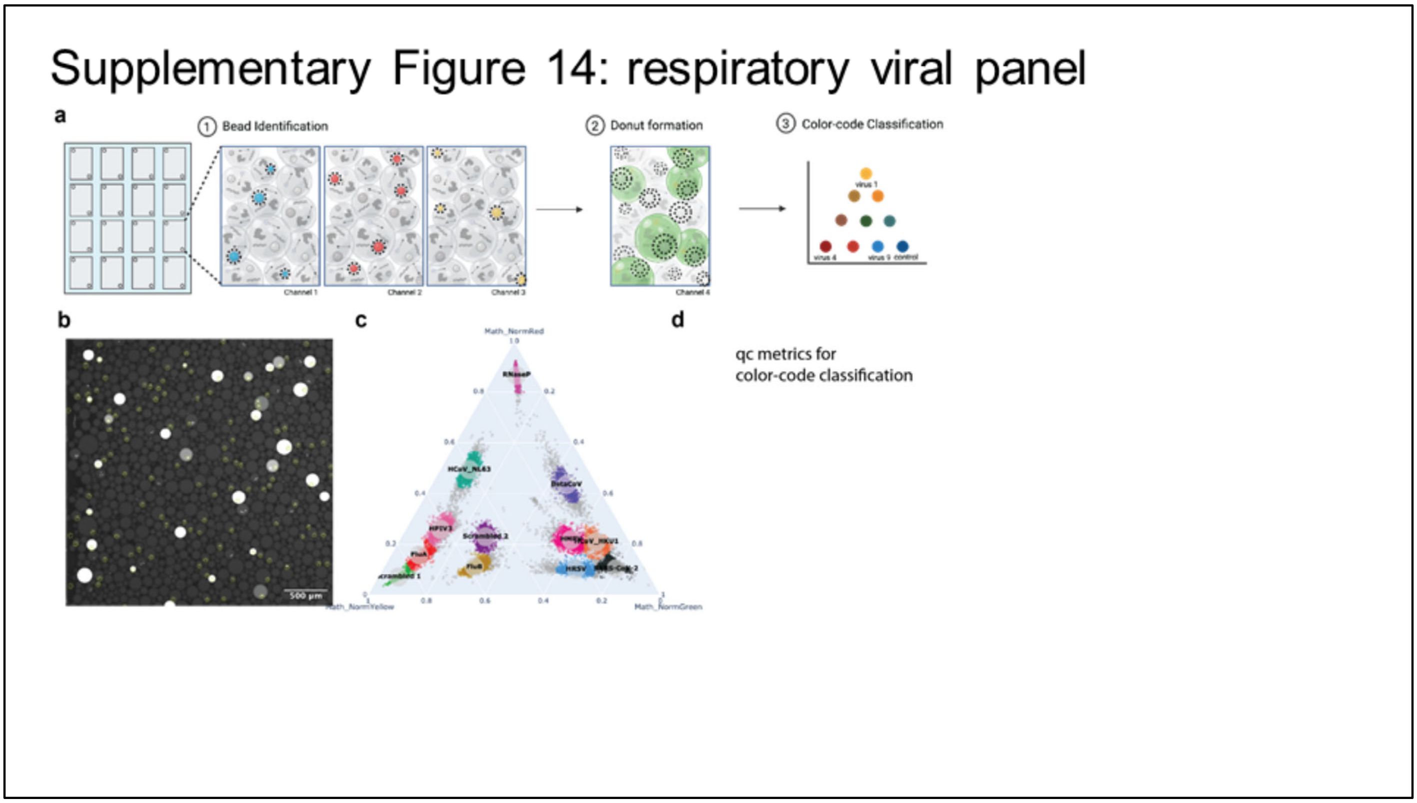


**Supplementary Figure 14. bbCARMEN setup and image analysis for bead classification with RVP. a**, schematic of bbCARMEN reaction setup and image analysis pipeline that: 1) identifies beads within droplets by drawing a circle around a single bead in droplet, 2) draws an additional circle surrounding the identified bead for FAM signal analysis, and 3) bead color-codes are characterized and classified. **b**, image of bead identification. **c**, ternary plot characterizing the 12 color codes that make up the RVP bead pool.

### Supplementary Figure 15: Characterization of RVP fluorescence across bead and lane replicates

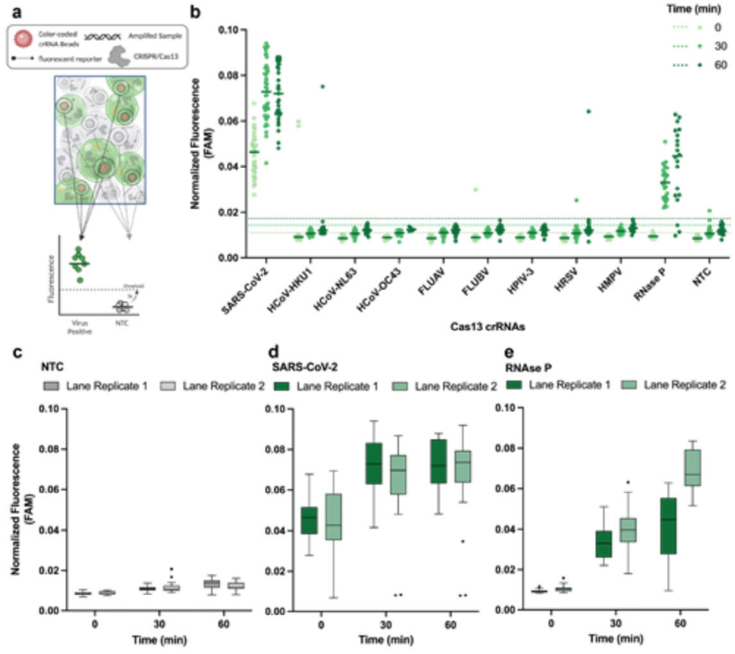


**Supplementary Figure 15. Characterization of SARS-CoV-2 FAM fluorescence signal across all bead and lane replicates. a**, Schematic of fluorescence values derived from signals within the donuts formed around a bead within a droplet. **b**, FAM fluorescence within donuts of each color-coded crRNA bead population that make up RVP at 0, 30, and 60 min post-reaction initiation. Bar at median fluorescence. Thresholds shown as dashed lines at each time point calculated as 3x the standard deviation of the NTC. **c-e**, Tukey box and whiskers plot of fluorescence values across lane replicates at 0, 30, and 60 min post-reaction initiation. Outliers represented as single points. **c**, NTC fluorescence. **d**, SARS-CoV-2 fluorescence. **e**, RNase P fluorescence.

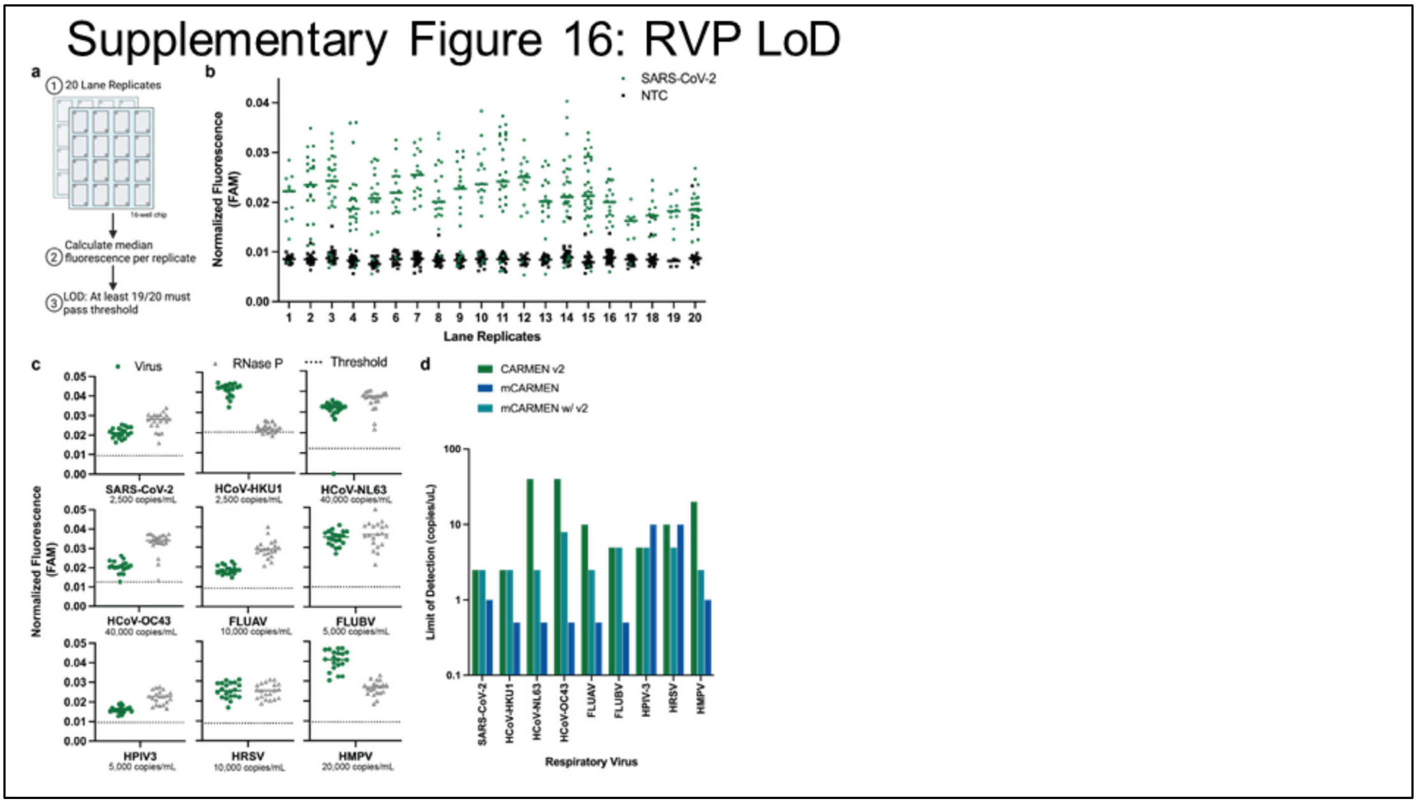


**Supplementary Figure 16. RVP limit of detection evaluation on bbCARMEN. a**, Schematic of LOD testing with bbCARMEN. **b**, Fluorescence of SARS-CoV-2, 2.5 copies/uL, and NTC for each of the 20 technical replicates. Individual points represent signal from a single droplet with a bar at median fluorescence. Green: SARS-CoV-2; Black: NTC. **c**, Median fluorescence (n=20) at the LOD for each of the 9 viruses on RVP as established by spiking synthetic RNA into negative control RNA. Green: virus; Gray: RNase P; Dashed line: Threshold derived from NTC. **d**, Comparison of RVP LODs across the CARMEN technologies. Green: bbCARMEN RVP assay; Teal: mCARMEN used w/ bbCARMEN RVP crRNA; Blue: mCARMEN used w/ mCARMEN RVP crRNA

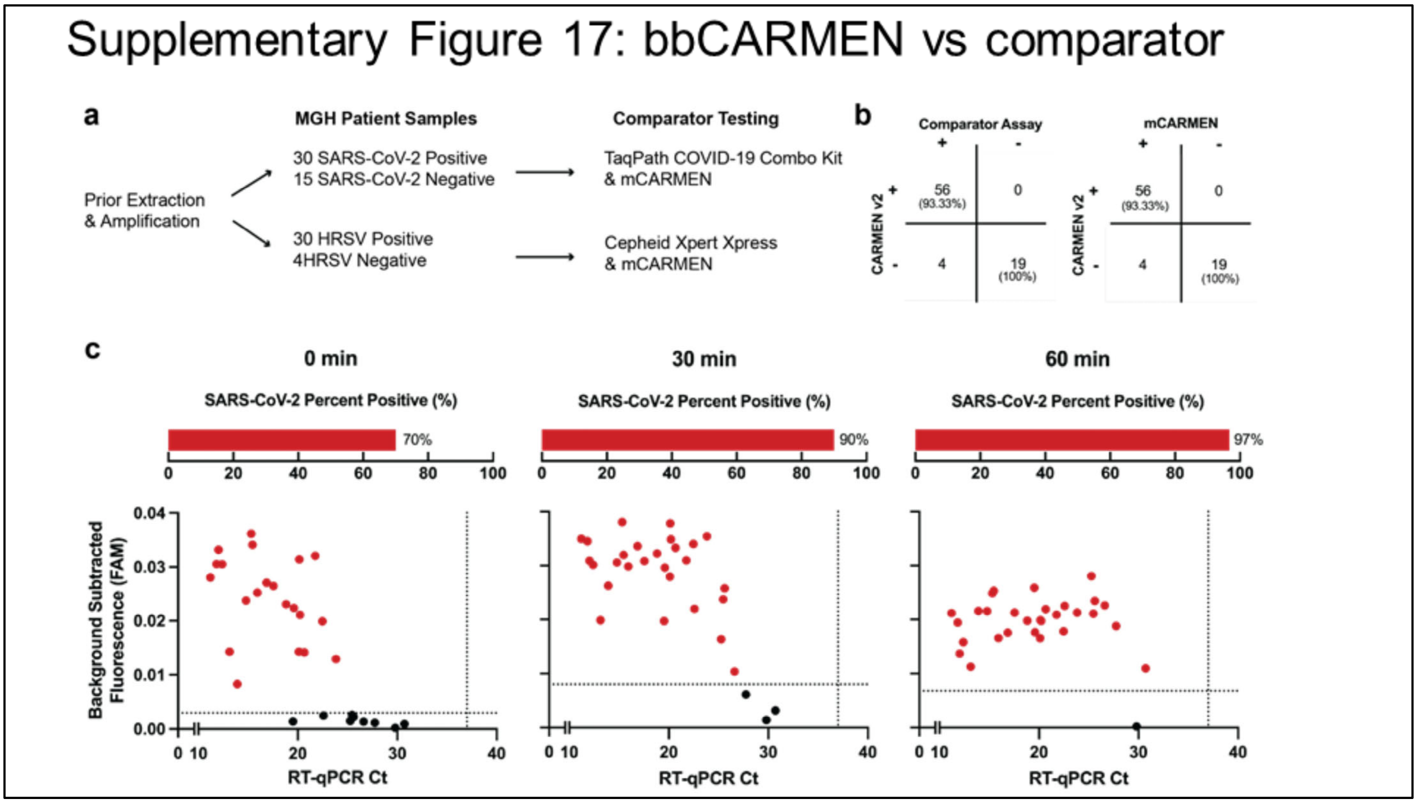


**Supplementary Figure 17. Comparison of bbCARMEN detection results to results collected from comparator assays. a**, Schematic of patient sample testing, with 79 patient samples tested, including 45 SARS-CoV-2 samples (30 positive, 15 negative) and 34 HRSV samples (30 positive, 4 negative). **b**, Concordance of bbCARMEN and bbCARMEN for 79 patient samples. **c**, Scatter plot of scaled normalized fluorescent values compared to viral Ct values detected by RT-qPCR at 0 min, 30 min, and 60 min timepoints for the positive SARS-CoV-2 samples.

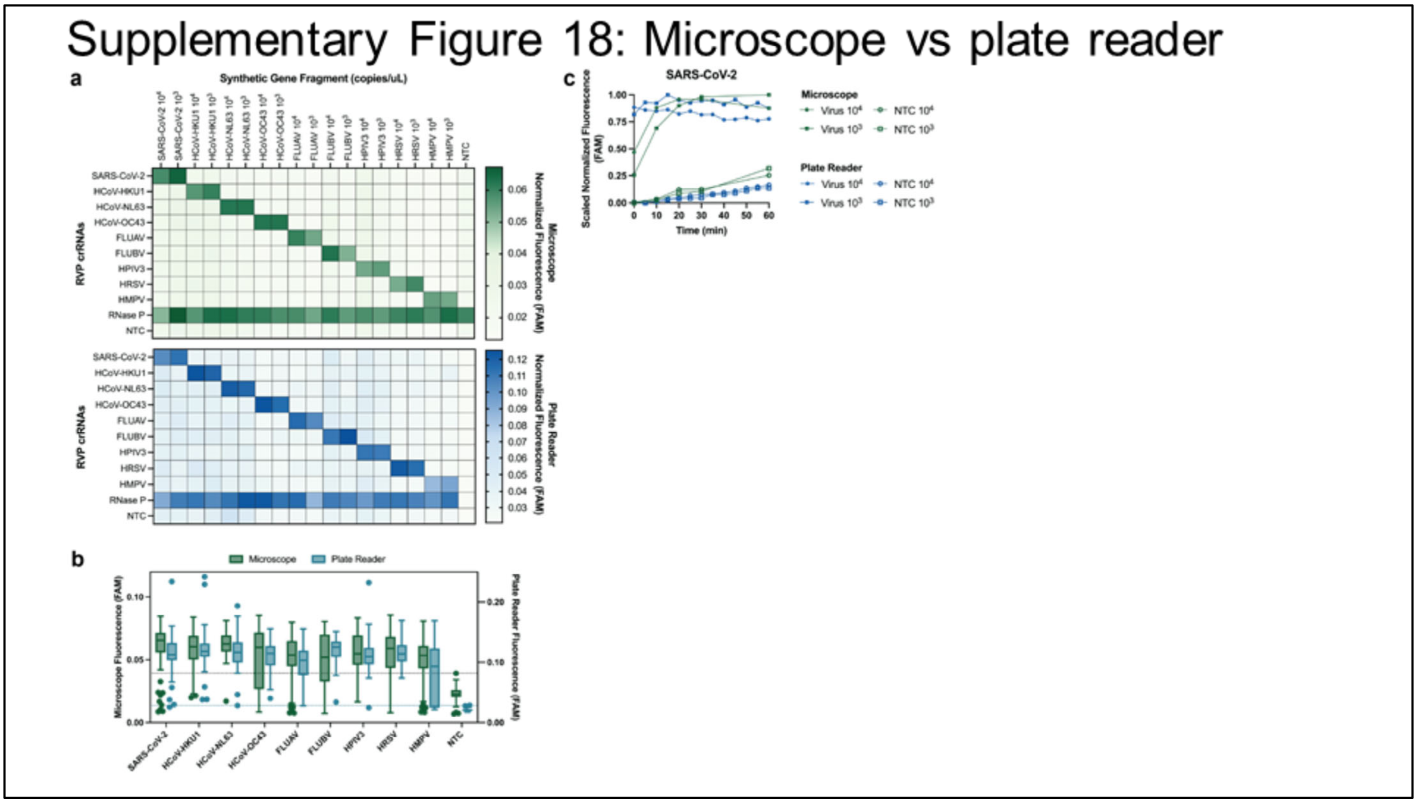


**Supplementary Figure 18. Comparison of bbCARMEN fluorescent results from two readout platforms. a**, Heatmaps at 60 min post-reaction initiation. **b**, Tukey box and whisker plots at 1e3 copies/uL 60 min post-reaction initiation; dashed line: 3 std dev above the median NTC. **c**, Kinetic curves of runs with 1e4 and 1e3 copies of SARS-CoV-2 on the microscope and plate reader-based platforms, including scrambled crRNA NTC values for each input sample.
